# Proliferation- and cytotoxic immune signatures identify chemotherapy-responsive bladder tumors in a molecular subtype dependent manner

**DOI:** 10.64898/2025.12.04.25341613

**Authors:** Aymeric Zadoroznyj, Pontus Eriksson, Carina Bernardo, Lena Tran, Carl-Adam Mattsson, Thomas W Flaig, Roland Seiler, Catherine M Tangen, Peter C Black, Seth P Lerner, David J McConkey, Mattias Höglund, Fredrik Liedberg, Gottfrid Sjödahl

**Author notes:** Corresponding author : Aymeric Zadoroznyj, Lund University, Medicon Village, Building 404, Scheelevägen 2, SE-223 81 Lund, Sweden.

## Abstract

Neoadjuvant cisplatin-based chemotherapy (NAC) followed by radical cystectomy (RC) has been the standard care for muscle-invasive bladder cancer (MIBC) for two decades. One third of NAC-treated patients achieve pathologic complete response (pT0N0), a proxy for improved survival after RC. Predicting response already at transurethral resection of bladder tumor (TUR-BT) would enable selective use of NAC, minimizing unnecessary toxicity and exposure to ineffective therapy. We aimed to identify tumor mRNAs predicting response across multiple transcriptomic studies, prioritizing subsequent biomarkers for validation.

Three NAC-treated and one RC-only cohort with tumor transcriptomic profiles were included. Differential mRNA-expression analysis and subtype classification according to the Lund Taxonomy were performed. Within each cohort and molecular subtype, genes were ranked by differential expression, and ranks were integrated across cohorts into a meta rank-score. Its predictive value for treatment response was assessed by resampling. Survival associations of top genes in the NAC- and RC-only cohorts were compared to select candidate biomarkers for protein level validation.

Proliferation/late cell-cycle gene expression predicted NAC response within the Urothelial-like subtype and expression of cytotoxic T- and NK-cell-related genes predicted response in Basal/Squamous tumors. These findings were validated by immunostainings for CCNB1 and NKG7, respectively.

This integrative framework suggests a complex picture in which two main NAC-predictive signals, the proliferation and the T/NK-cell signatures, identify responders in a subtype dependent manner. Meta rank-scores additionally suggest new candidate biomarkers in each Lund Taxonomy subtype. The applied framework can be updated as new datasets become available, providing dynamic exploration of NAC-predictive biomarkers in MIBC.

## Introduction

Cisplatin-based neoadjuvant chemotherapy (NAC) prior to radical cystectomy (RC) provides a modest but significant survival benefit of 5-8% for patients with muscle-invasive bladder cancer (MIBC) [1, 2]. The most commonly used NAC regimens are dose-dense MVAC, (methotrexate, vinblastine, doxorubicin and cisplatin) and GC (gemcitabine and cisplatin), which may be efficiently combined with immune checkpoint inhibition [3]. Although the benefit of NAC is mediated by treating micrometastatic disease, pathologic complete response (pCR) in the bladder and excised lymph-nodes (pT0N0) is strongly associated with favorable long-term survival [4–6]. In non-responders on the other hand, outcomes might even worsen due to NAC by further delaying RC [7, 8]. This underscores the need for predictive biomarkers to identify patients most likely to respond, and reserve NAC for those patients.

Several tumor features have been associated with NAC response, including molecular subtypes [9–11], genomic alterations in DNA damage response genes such as ERCC2 and BRCA2 [9, 12], immune infiltration [13–15], and histological architecture [16]. However, validation of these biomarkers and understanding of their relationship with subtype classification is still unclear [17, 18], and no molecular predictors are currently used for clinical decision-making. We hypothesize that two main shortcomings have limited progress in this field. First, MIBC has until now been treated as one disease, without considering that molecular subtypes may have different response and resistance mechanisms. Second, the reliance on single cohorts with small sample sizes lead to high false discovery rate and low concordance between study results.

Here, we combine the three largest transcriptomic datasets with MIBC patients treated with NAC, to explore gene expression patterns and candidate biomarkers predictive of pCR within each molecular subtype.

## Materials and methods

### Identification and acquisition of datasets

The literature was screened to identify cohorts of patients with MIBC receiving cisplatin-based NAC followed by RC. Cohort inclusion criteria were as follows; transcriptomic data for at least 100 patients to enable subtype-specific analysis, available response status (ypTN) following at least 2 cycles of NAC. By December 2024, three patient cohorts were selected: one in-house dataset, the ‘Lund’ Chemo-cohort [10] (*n*=149), and two external datasets, the ‘SWOG S1314’ cohort (*n*=227) [19, 20] and the ‘Seiler2017’ cohort (*n*=262) [21]. Within the SWOG S1314 cohort, 64 patients were excluded, 45 due to absence of transcriptomic data and 19 due to non-evaluable pathologic response (e.g. no surgery or ypTX). From the Seiler2017 cohort, 3 patients without information about ypTN status were excluded. The ‘RC-only’ cohort, a Swedish population-based consecutive RC cohort not receiving peri-operative chemotherapy (*n*=186) [22, 23], was used as comparison to disentangle prognostic from predictive associations.

### Microarray data preprocessing

RNA expression data for the Lund Chemo- and RC-only cohorts was generated on the Affymetrix Human Gene 1.0 ST microarray platform and are available from the Gene Expression Omnibus data repository under accession numbers GSE169455 [10] and GSE83586 [22]. Raw data were RMA (Robust Multichip Average) preprocessed and ComBat adjusted to account for differences between labeling batches. Probe sets were then annotated using the BrainArray V25 based on Gencode 36 and summarized at the gene level, resulting in 17,953 unique genes. RNA expression data from Seiler and colleagues [21] generated on the Affymetrix Human Exon 1.0 ST microarray platform was downloaded from the Gene Expression Omnibus data repository under accession number GSE87304. RNA expression data from the SWOG S1314 trial was generated on the Affymetrix Human Genome U133 Plus 2.0 Array platform and provided by the Southwest Oncology Group. The Seiler2017 and SWOG S1314 datasets were processed in the same way as the Lund cohorts, resulting in 18,869 and 15,987 unique genes respectively. Genes present in all cohorts were retained for analysis, corresponding to 15,757 genes.

### Lund and consensus subtype classification

Lund molecular subtype classification of all samples was performed using the latest version of the Lund Taxonomy (LundTax) classifier, available through the LundTax2023Classifier R package [24]. Tumors were grouped into the five main molecular subtypes: Urothelial-like (Uro), Genomically unstable (GU), Basal/Squamous (Ba/Sq), Mesenchymal-like (Mes), and Small-cell/Neuroendocrine-like (Sc/Ne). Consensus molecular subtype classification of MIBC was performed using the R package consensusMIBC [25]. Tumors were classified into six molecular subtypes: luminal papillary (LumP), luminal non-specified (LumNS), luminal unstable (LumU), stroma-rich, Ba/Sq, and neuroendocrine-like (Ne-like).

### Meta rank-score calculation and evaluation

In each dataset, differential expression analysis was performed between patients with and without pCR (ypT0N0) using the R package ‘Limma’. Volcano plots were generated using GraphPad Prism (v10.4.1). For each dataset, genes were ranked from 1, most associated with response, to 15,757, most associated with non-response, based on the moderated t-statistic. To generate a combined ranking across the three NAC-treated cohorts, ranks were weighted by the total number of significant genes in each cohort (adjusted p<0.05), giving greater weight to cohorts with the higher discriminatory power. The weighted rank sum for each gene across the three cohorts was then used to obtain the meta rank-score ranging from 122.3 for the gene most strongly associated with treatment response to 35,336.2 for the gene most strongly associated with non-response. Meta rank-score calculation was performed on the full cohort but also separately for the most prevalent molecular subtype categories; Uro, GU, Ba/Sq, luminal subtypes (Uro or GU), non-luminal subtype (Ba/Sq, Mes, or Sc/Ne), and for treatment modality categories MVAC and GC. Genes were also separately ranked by differential expression analysis in the Lund RC-only cohort. The same analyses were performed using the MIBC consensus classification. Resampling analysis was performed using identical meta rank-scores for 100 permutations in each dataset with random pCR label assignment. The extremes of the permuted meta rank-score distributions were then compared to the observed meta rank-scores. At each position of the distribution, a NAC response signal was defined by a one-sided empirical p<0.05, i.e. if the observed meta rank-score was more extreme than the fifth most extreme permutation.

### GO term analyses, GSEA, PPI analysis and proliferation score analysis

Gene ontology (GO) enrichment analysis of biological processes was performed using the ShinyGO tool (v0.82) [26]. Terms overrepresented in the 1% or 5% of genes with the highest or lowest meta rank-score were identified for each molecular subtype and treatment category using all common gene as background. Gene set enrichment analysis (GSEA) was performed on the full cohort, and separately for molecular subtype and treatment categories, using GSEA (v4.3.2), scanning H (hallmark), C2 (curated), and C6 (oncogenic) MSigDB gene sets [27], with genes pre-ranked according to the meta rank-score. Protein-Protein Interaction (PPI) networks were generated using Cytoscape (v3.10.3) and StringApp (v 2.2.0) [28] on the 1% of genes with the highest or lowest meta rank-score in each subtype. The full STRING network, with a confidence score of 0.7, and a minimum of 5 interactors was used for the analysis. The Proliferation score was calculated using the ‘LundTaxR’ package (https://github.com/mattssca/LundTaxR).

### Immunohistochemical validation

Immunohistochemical (IHC) validation was performed on tissue microarrays (TMA) from the Lund Chemo and RC-only cohorts described previously [10, 22]. Briefly, two 1.0 mm tissue cores were obtained per tumor. Cores were placed at representative, tumor-rich areas of the same formalin-fixed paraffin embedded tissue block that was used for RNA extraction. TMA-sections (3 μm) were stained with primary antibodies for SMCO4 (Polyclonal, Proteintech), CYP4Z1 (STJ92604, St John’s Laboratory), CD55 (EPR6689, Abcam), ALDH1A1 (EP1933Y, Abcam), NKG7 (F4V5I, Cell signaling) and CCNB1 (Y106, LifeSpan-BioSciences). Sections were stained on the automated Ventana DISCOVERY ULTRA (Ventana Medical Systems Inc, Tucson, AZ) instrument using DAB for detection, scanned (S60, Hamamatsu), and evaluated as digital images with PathXL Tutor (Cirdan Imaging Limited, UK). Staining was assessed according to the intensity of labelling (score 0-3) and/or the percentage of labelled cells (in 10% increments for cancer cells or stromal cells), depending on each protein’s cellular expression pattern. For proteins positive in both cancer cells and stromal cell types, scores for the two compartments were recorded separately. For each case, the mean value for the two TMA cores was used in the analyses.

### Statistical analyses

Statistical analyses and visualizations were performed using the R software (v4.3.2) or the GraphPad Prism software (v10.4.1). Univariate Cox proportional hazards regression was performed for the Lund Chemo- and RC-only cohorts using the R package ’survminer’. Normality tests and two-sided non-parametric tests with a significance threshold of 0.05 were used for validation of biomarkers by IHC.

## Results

### Response to NAC is impacted by tumor molecular subtype

We analyzed RNA expression data and pathologic response outcomes from three bladder cancer cohorts treated with MVAC or GC; the ‘Lund’ (*n*=149), the ‘SWOG S1314’ (*n*=163) and the ‘Seiler2017’ (*n*=259) cohorts (**Figure 1A**). In each cohort, patients with pCR were compared with patients who had residual disease in the cystectomy specimen. An additional ‘RC-only’ cohort (*n*=186), composed of patients who underwent RC without NAC, was used as a control. A schematic overview of the analytical pipeline is shown in **Figure 1B**.

**Figure 1.**
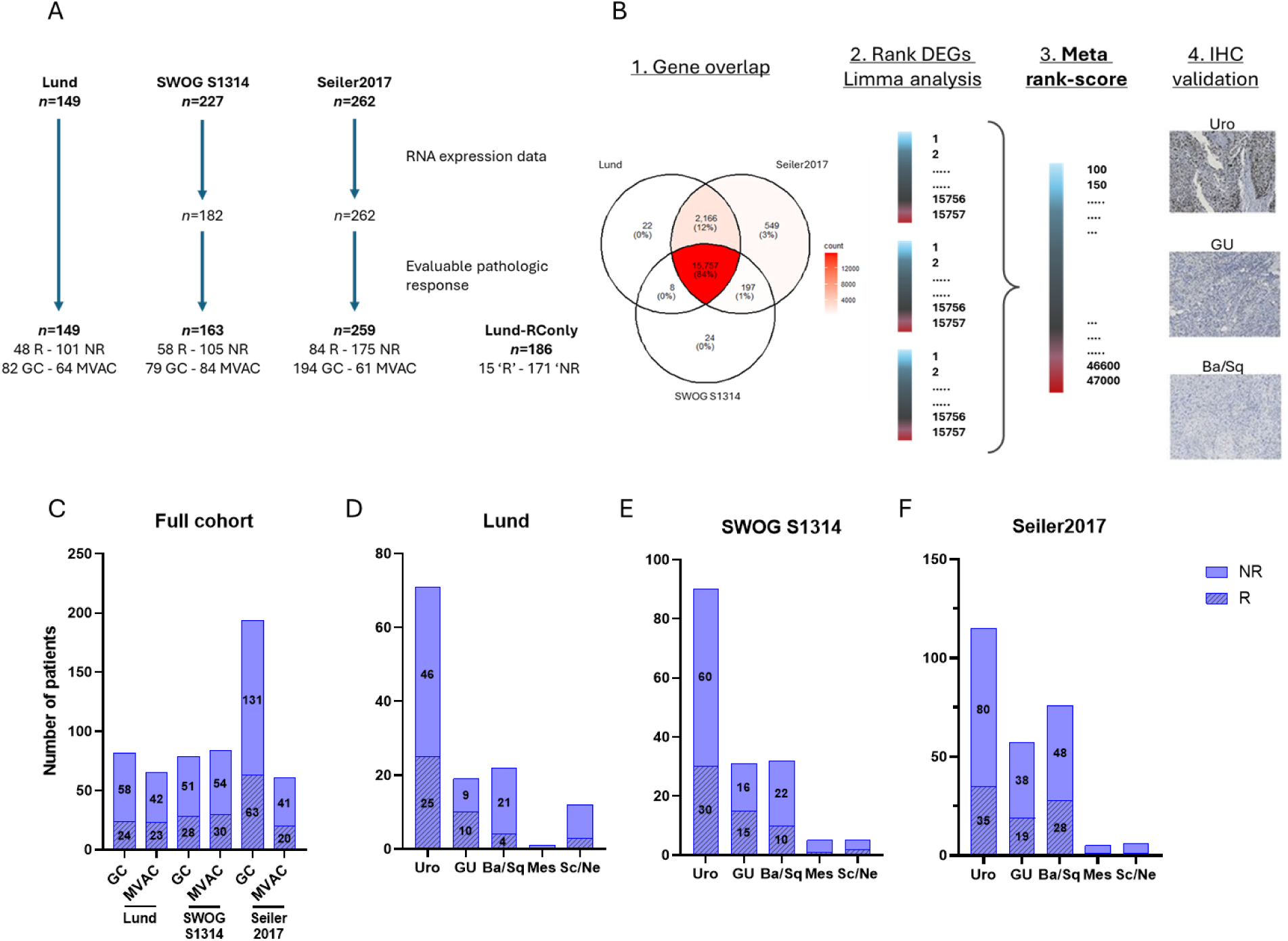
Classification of bladder cancer by subtype highlights discrepancies in response to NAC. A. Schematic representation of the three NAC-treated cohorts and patient inclusion criteria. B. Description of the research pipeline. Only genes common to all cohorts were selected. Limma differential expression analysis was performed on the full cohort and by molecular subtypes, and the results were combined to obtain a meta *rank-score. The most promising biomarkers identified using the meta rank-score were selected and validated by immunohistochemistry. C-F. Comparison of treatment response between the full cohort (C) and among the 5 molecular subtypes within each cohort: Lund (D), SWOG S1314 (E) and Seiler2017 (F). R: responder defined as pCR; NR: non-responder*.

Each patient cohort was classified into five main molecular subtypes according to the Lund Taxonomy, where the Uro subtype can be further divided into three minor subtypes (UroA, B and C) (**Figure S1**-**3**). Treatment response, defined as pCR (ypT0N0), was stratified by NAC-regimen (GC and MVAC) (**Figure 1C**) and by molecular subtype within each cohort (**Figure 1D-F**). Subtype specific differences were most evident in the Lund and SWOG S1314 cohorts, where patients with the GU subtype responded better than those with a Ba/Sq subtype (**Figure 1D-E**). This pattern was not observed in the Seiler2017 cohort (**Figure 1F**). The most prevalent Uro subtype had a consistent intermediate response rate across all cohorts, while the less prevalent Mes and Sc/Ne subtypes were too infrequent for analysis.

### Meta rank-score analysis highlights subtype-dependent biomarkers of NAC sensitivity

Differential expression analysis by pCR status in each cohort identified 817, 566, and 1,084 differentially expressed genes (DEGs) in the Lund, SWOG S1314 and Seiler2017 cohorts, respectively (**Figure 2A**). The same analysis was performed within each molecular subtype for all NAC cohorts (**Figure 2B-D**) and for MVAC versus GC regimen (Figure S4). Similar analyses were performed per subtype within the Lund RC-only control cohort (**Figure S5**). Genes consistently detected across the three datasets were ranked according to their association with pCR, yielding a weighted meta rank-score. Lower meta rank-scores corresponded to genes positively associated with response, whereas higher scores indicated association with non-response. Meta rank-scores for all 15,757 genes are presented in **Table S1.** The meta rank-score exploration identified *KBTBD4*, *C17orf97*, *RACGAP1*, *C9orf64* and *SMCO4* as genes most associated with NAC sensitivity in the full cohorts, with subtype-specific top genes including *SMCO4*, *FOXM1*, *KNSTRN*, *CDKN3* and *KIF20A* in Uro; *MAFG*, *SHMT1*, *LHX4*, *DNER* and *RPH3AL* in GU; and *NKG7*, *NOXRED*, *GZMB*, *GZMH* and *TCN2* in Ba/Sq (**Table 1**). Two clear patterns emerged from the analysis: proliferation genes (e.g. *FOXM1, CDKN3, and KIF20A*) were associated with NAC sensitivity in the Uro subtype, whereas genes expressed by cytotoxic immune cells (e.g. *GZMB*, *GZMH* and *NKG7*) predicted a favorable response in the Ba/Sq subtype. In contrast, the top genes associated with non-response did not reveal any consistent biological theme in any of the subtypes. Across all analyses, the lowest meta rank-score was observed in the Uro subtype for the gene *SMCO4* with ranks 1, 49, and 96 across the three cohorts. Conversely, the highest meta rank-score was obtained in the Ba/Sq subtype for *SVIL*, which encodes the actin regulatory protein supervillin [29]. The top genes associated with response and non-response in each subtype are highlighted in the corresponding volcano plots for each NAC cohort (**Figure 2 A-D**).Taken together, comparison of the meta rank-scores in the full cohort and the subtype-informed analyses showed increased biological specificity when accounting for molecular subtypes. Consequently, in the full cohort, the proliferation and immune-cytotoxicity signals identified in Uro and Ba/Sq subtypes, respectively, were diluted and masked by mixed signatures and other genes. Importantly, the top-ranked genes in each subtype showed no association with pCR in the Lund RC-only cohort (i.e., pT0N0 after TUR-BT alone), suggesting that these signals reflect NAC response rather than identifying patients with low disease burden (**Table 1** and **Figure S5**).

**Figure 2.**
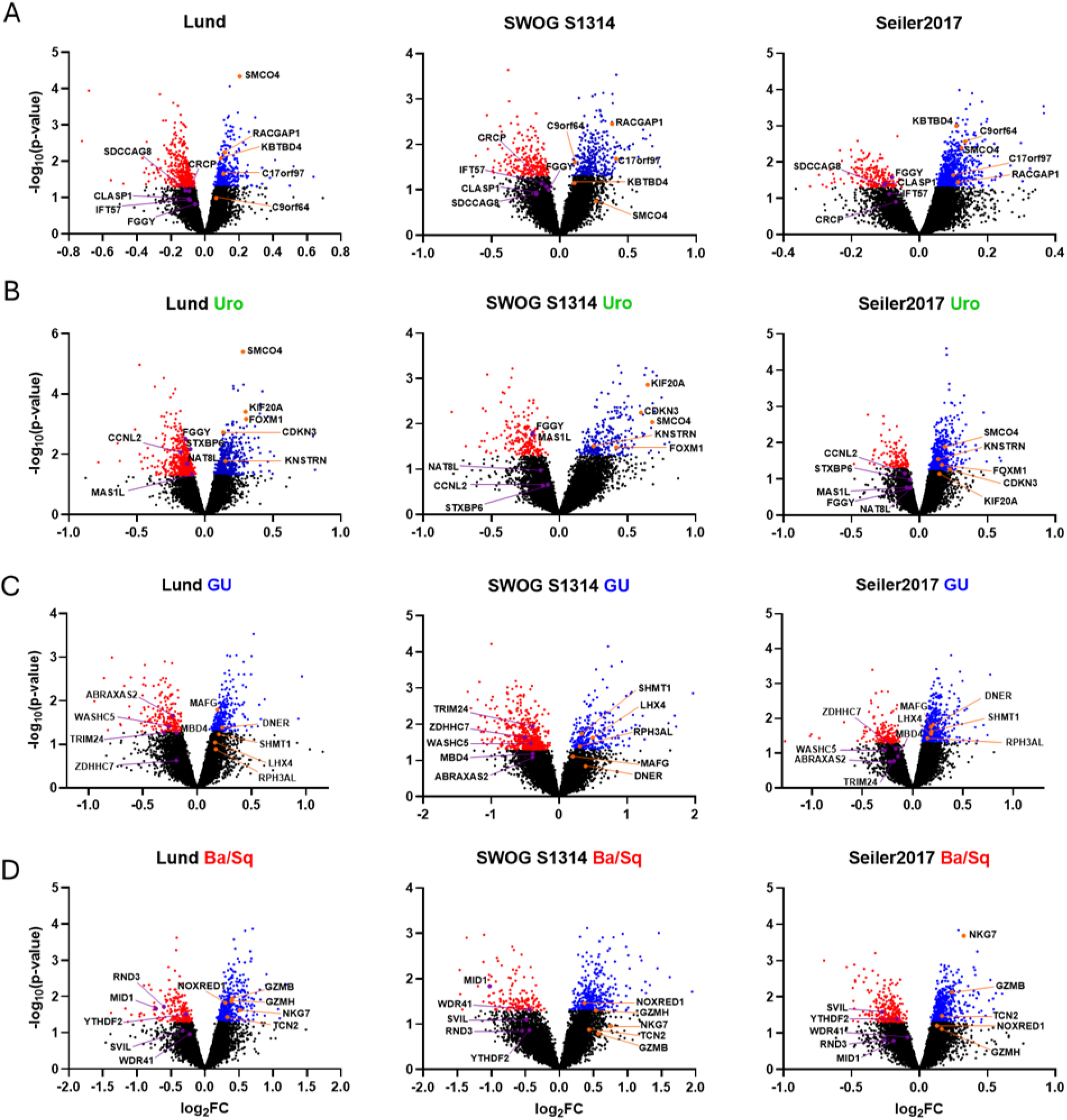
Differential expression analysis by treatment response in each cohort and in three molecular subtypes. A-D. Volcano plots representing the differential expression analyses performed on the NAC-treated full cohort (A) and the three major molecular subtypes: Uro (B), GU (C) and Ba/Sq (D). Genes with statistical significance (p≤0.05) are marked in red for downregulation and blue for upregulation, and the 5 genes with the highest and lowest meta rank-score in each volcano plot are highlighted.

**Table 1.** Top 5 genes with the highest and lowest meta rank-score in the full cohort and in each major subtype. *For each gene, the rank in each cohort and their exact* meta rank-score *is described. The Lund RC-only rank is shown as a control*.

| Subset | Direction | Top 5 Genes | Rank Lund | Rank SWOG S1314 | Rank Seiler2017 | Meta rank-score | Rank Lund RC-only |
| --- | --- | --- | --- | --- | --- | --- | --- |
| Full cohorts | Response | *KBTBD4* | 43 | 494 | 13 | **303.4** | 12,735 |
|  |  | *C17orf97* | 118 | 149 | 334 | **500.7** | 12,889 |
|  |  | *RACGAP1* | 32 | 19 | 519 | **553** | 1,871 |
|  |  | *C9orf64* | 672 | 183 | 28 | **630** | 1,873 |
|  |  | *SMCO4* | 1 | 1,291 | 48 | **722.8** | 9,423 |
|  | Non-response | *FGGY* | 14,515 | 15,342 | 15,674 | **34,624.5** | 12,344 |
|  |  | *CLASP1* | 14,820 | 15,177 | 15,602 | **34,696.2** | 9,568 |
|  |  | *CRCP* | 15,256 | 15,262 | 15,235 | **34,702.2** | 1,826 |
|  |  | *IFT57* | 14,878 | 15,402 | 15,467 | **34,722.4** | 14,714 |
|  |  | *SDCCAG8* | 15,272 | 15,119 | 15,628 | **35,032.6** | 5,971 |
| Uro | Response | *SMCO4* | 1 | 49 | 96 | **122.3** | 5,363 |
|  |  | *FOXM1* | 12 | 172 | 307 | **405.9** | 8,001 |
|  |  | *KNSTRN* | 175 | 156 | 199 | **412.4** | 8,219 |
|  |  | *CDKN3* | 26 | 32 | 417 | **453.3** | 6,850 |
|  |  | *KIF20A* | 9 | 9 | 574 | **585.5** | 2,801 |
|  | Non-response | *NAT8L* | 15,527 | 15,243 | 15,154 | **34,815.5** | 12,781 |
|  |  | *STXBP6* | 15,672 | 14,457 | 15,459 | **34,819.4** | 14,288 |
|  |  | *MAS1L* | 15,261 | 15,701 | 15,178 | **34,878.2** | 8,625 |
|  |  | *CCNL2* | 15,655 | 14,369 | 15,597 | **34,898.7** | 1,231 |
|  |  | *FGGY* | 15,717 | 15,707 | 15,164 | **35,211.0** | 10,312 |
| GU | Response | *MAFG* | 60 | 444 | 182 | **459.1** | 7,222 |
|  |  | *SHMT1* | 278 | 106 | 259 | **523.9** | 8,668 |
|  |  | *LHX4* | 484 | 136 | 299 | **734.8** | 8,167 |
|  |  | *DNER* | 182 | 852 | 154 | **736.0** | 10,106 |
|  |  | *RPH3AL* | 755 | 234 | 413 | **1104.2** | 10,916 |
|  | Non-response | *ZDHHC7* | 14,455 | 15,527 | 15,508 | **34,509.9** | 1,652 |
|  |  | *ABRAXAS2* | 15,694 | 14,898 | 15,071 | **34,678.3** | 7,297 |
|  |  | *MBD4* | 15,566 | 15,079 | 15,289 | **34,894.3** | 6,844 |
|  |  | *TRIM24* | 15,569 | 15,650 | 15,035 | **34,940.7** | 9,625 |
|  |  | *WASHC5* | 15,619 | 15,410 | 15,462 | **35,280.1** | 2,902 |
| Ba/Sq | Response | *NKG7* | 179 | 886 | 2 | **599.5** | 8,462 |
|  |  | *NOXRED1* | 110 | 229 | 635 | **837.5** | 12,770 |
|  |  | *GZMB* | 104 | 1,366 | 49 | **840.6** | 5,930 |
|  |  | *GZMH* | 90 | 342 | 800 | **1046.4** | 6,250 |
|  |  | *TCN2* | 260 | 1,073 | 323 | **1079.2** | 9,345 |
|  | Non-response | *RND3* | 15,712 | 15,143 | 14,938 | **34,686.8** | 1,526 |
|  |  | *WDR41* | 15,261 | 15,629 | 15,091 | **34,753.6** | 946 |
|  |  | *MID1* | 15,707 | 15,730 | 14,893 | **34,944.5** | 6,188 |
|  |  | *YTHDF2* | 15,669 | 15,167 | 15,584 | **35,312.9** | 3,269 |
|  |  | *SVIL* | 15,404 | 15,462 | 15,653 | **35,336.2** | 11,540 |

### Broad proliferation and immune cytotoxicity signatures have strong subtype-dependent impact on NAC-response

To systematically investigate the biological pathways underlying NAC response, we performed gene ontology (GO) analysis using the 1% or 5% of genes with the lowest and highest meta rank-score, both for the full cohort and within each major subtype (Uro, GU and Ba/Sq) (**Figure 3A-D** and **Figure S6A-D**).Genes associated with non-response to NAC, (highest meta rank-scores), showed no consistent biological patterns, with only modest enrichment for cytoskeleton organization in the full cohort and within the Ba/Sq subtype. However, for genes associated with treatment sensitivity, a strong enrichment of cell cycle and cell proliferation related processes was observed (**Figure 3A**). Similar proliferation-related genes were further enriched in the Uro subtype, with approximately doubled levels of enrichment (top 20 terms: ∼2-8 fold in the full cohort versus ∼4-20 fold in Uro) and markedly stronger false discovery rate (FDR) adjusted p-values (10^-3^-10^-6^ for full cohort versus10^-8^-10^-12^ for Uro, **Figure 3B**). We observed no statistically robust enrichments related to response in the GU subtype (**Figure 3C**), whereas the Ba/Sq subtype showed strong enrichment (top 20 terms: ∼2-4-fold enrichment and FDR-adjusted p=10^-20^-10^-40^) for immunity- and cytotoxicity-related processes (**Figure 3D**). These subtype-specific enrichments were maintained when analyzing the top 1% and the top 5% of genes (**Figure S6**). These results were further validated by GSEA analysis: 128 and 147 gene sets were significantly enriched (FDR≤0.05) in the Uro and Ba/Sq subtypes, respectively, corresponding to cell proliferation and cytotoxic immune response, (**Figure S7**) whereas no significant enrichments were identified in GU. To further investigate response-associated biology, we used STRING to test if any protein interaction networks are differentially regulated depending on the meta rank-score (**Figure 3E-F)**. Consistent with the ontology analysis, no response-related networks were detected in the GU subtype. As expected, however, the response-associated genes in the Uro subtype formed a dense proliferation-related network with discernible substructures relating to nucleosome and kinetochore structures as well as homologous recombination repair (**Figure 3E**). An interactome related to T/NK-cell immunity was also seen for response-associated genes in the Ba/Sq subtype (**Figure 3F**), although it was not as dense as the proliferation network in the Uro subtype. The network captured various aspects of the T cell interactome, e.g. T-cell receptor components (*CD3E*, *CD8A*), *IFNG*, cytotoxic granule proteins (*PRF1*, *GZMH*, *NKG7*), and immune checkpoints (*CD247*, *CTLA4*, *IDO1*). In the full cohort, the interactome profile was primarily driven by the Uro subtype, while a more limited overlap was noted with the Ba/Sq subtype (**Figure S8**).

**Figure 3.**
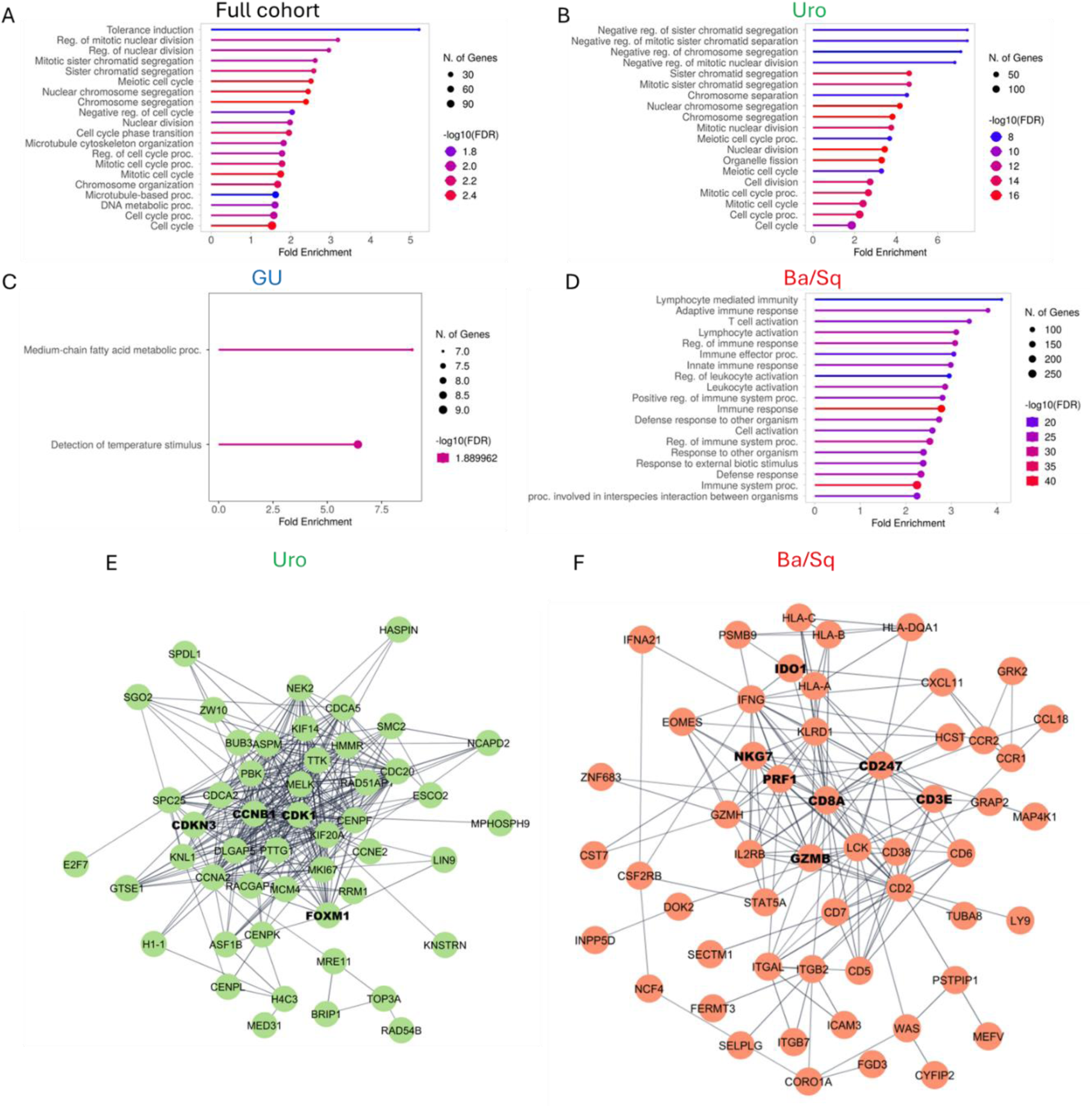
Cellular processes related to treatment response are specific to molecular subtypes. A-D. GO term analysis using ‘biological processes’, from the 5% of genes with the lowest meta rank-score in the full cohort (A) and major molecular subtypes: Uro (B), GU (C) and Ba/Sq (D). At most, the top 20 terms with the highest fold enrichment are described. E-F. Major protein-protein interaction observed in the top 1% of genes with the lowest meta rank-score in the Uro (E) and Ba/Sq (F) subtypes using STRING.

Similar to the GO analysis, genes associated with non-response did not form coherent networks suggesting that resistance mechanisms may be pleiotropic and less subtype-specific than the positive determinants of NAC sensitivity.

### MIBC Consensus classification identifies the same subtype dependent signatures

Differential expression analysis and meta rank-score calculation were also conducted for each subtype of the MIBC consensus molecular classification and for a combination of LumNS and LumP (corresponding to the LundTax Uro subtype) (**Table S2 and figure S9**). GO analysis was then performed on the top 5% of genes with the lowest meta rank-scores within each group (**Figure S10**). Consistent with previous findings, the Ba/Sq subtype showed enrichment of immune- and cytotoxicity-related processes among genes associated with treatment response (**Figure S10A**), though with weaker p-values than those observed in the LundTax classification (FDR-adjusted p = 10⁻² - 10⁻⁴). In the LumNS subtype, corresponding to a subset of the LundTax Uro subtype (**Figure S9**), response-associated genes were enriched for cell cycle and mitochondrial fission signatures (**Figure S10B**), again with weaker p-values than in the corresponding LundTax Uro subtype (FDR-adjusted p = 10⁻³ - 10⁻⁹). In LumNS and LumP combined, it is mainly cell cycle and proliferation signatures that are highlighted (**Figure S10C**). No statistically robust enrichments were detected for the LumP, LumU, or stroma-rich subtypes. Thus, the consensus classification supports our observations based on the LundTax classification, although with reduced statistical significance.

### Resampling-based analysis and pairwise cohort analysis confirm subtype specific predictive signals

Next, we assessed how informative response-related signals in the TUR-BT transcriptome are for predicting NAC response, and whether these signals are consistent across studies. For this purpose, we first investigated pairwise concordance of top differentially expressed genes across the three NAC cohorts (**Table S3**). We found that 79 of the top 500 response-associated genes in the Uro subtype overlapped between the Lund and the SWOG S1314 cohorts, with 40 of the overlapping genes being involved in cell proliferation. In contrast, the Seiler2017 cohort showed poor overlap with both the Lund and SWOG S1314 (23 and 28 genes, respectively) (**Figure 4A**). For the top 500 response-associated genes in the Ba/Sq subtype, we observed larger overlap between Lund and Seiler2017 cohorts (36 genes) than for SWOG S1314 with either Lund or Seiler2017 cohorts (17 and 16 genes, respectively) (**Figure 4B**). Among these 36 genes, 19 belong to an immune-related GO signature. These observations are supported by GO term analyses performed on the individual cohorts. Enrichment of cell proliferation signatures was observed in the Uro subtype in both the Lund and SWOG S1314 cohorts (**Figure S11A-B**), but not in the Seiler2017 cohort (**Figure S11C**).

**Figure 4.**
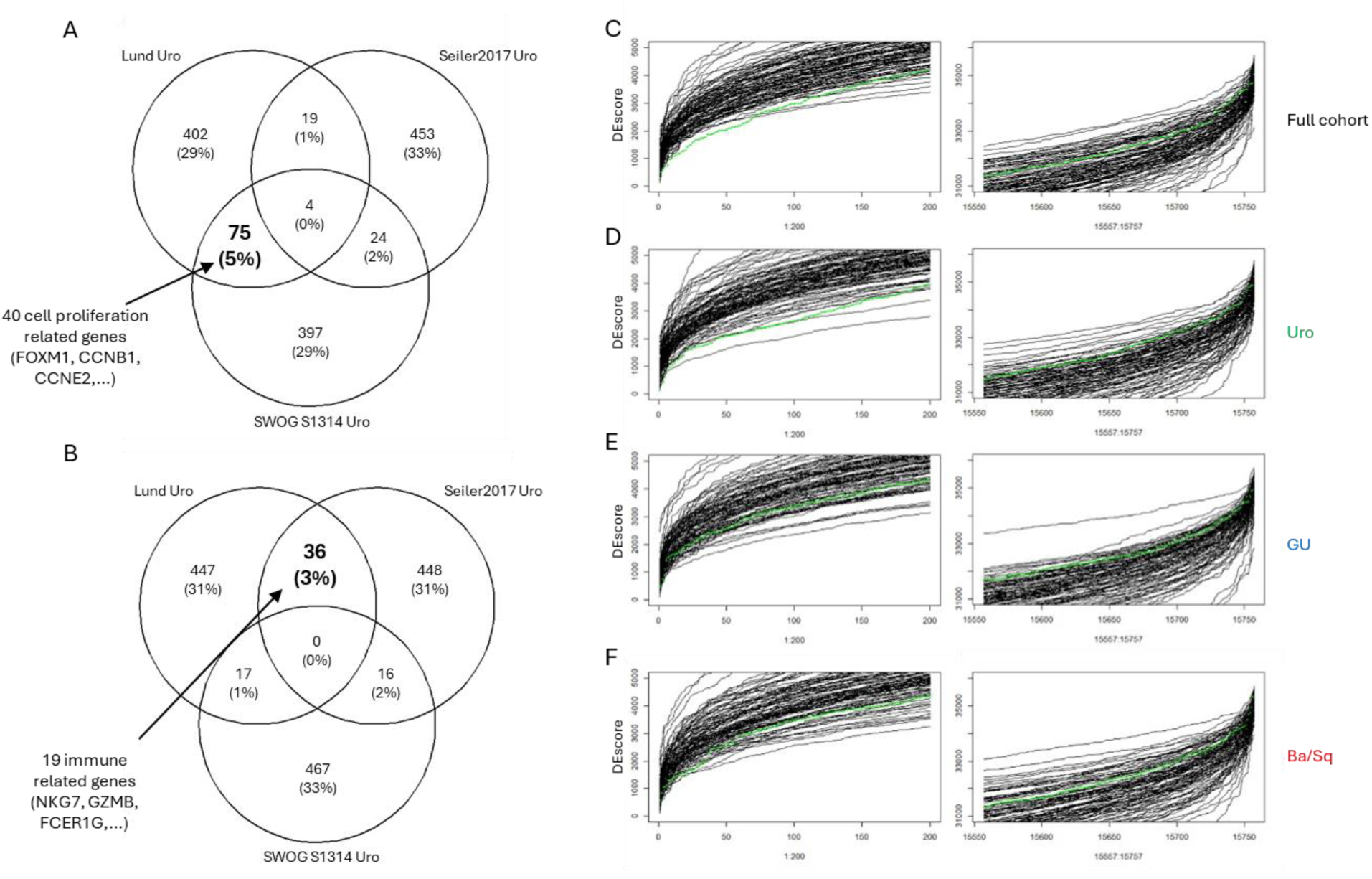
Meta rank-score resampling-based analysis highlights genes associated with treatment sensitivity. A-B: Venn diagram showing the number of genes associated with treatment sensitivity (500 genes with the lowest meta rank-score) that are shared across the different cohort in the Uro (A) and Ba/Sq (B) subtypes. C-F. Resampling-based analyses were performed for the full cohort (C) and in each major subtype (Uro, D; Ba/Sq, E; GU F). Line plots show meta rank-score distributions across the 200 genes with the lowest (left), and highest (right) scores. The observed meta rank-score distribution is shown in green and distributions from resampled data in black.

Conversely, enrichment of immune-related signatures was observed in the Ba/Sq subtype in the Lund and Seiler2017 cohorts (**Figure S11D-E**), however not in the SWOG S1314 cohort.

Next, we identified the response associated genes stratified by chemotherapy regimen (GC vs. MVAC). As was observed in the Uro subtype, proliferation signatures were enriched in responders also among patients treated with MVAC (top 20 terms: 2-4-fold enrichment and FDR-adjusted p=10^-10^-10^-16^) (**Figure S12A**) but not among those treated with GC (**Figure S12B**). To disentangle if the predictive effect of proliferation depended on Uro subtype, MVAC regimen, or both, we analyzed a proliferation score in responders and non-responders of the various subsets in each cohort separately (**Figure S13**). In the Lund and SWOG S1314 cohorts, the association between proliferation and response was limited to the Uro subtype, within which it was statistically significant only in the MVAC treated subset (**Figure S13A-D**). In the Seiler cohort, proliferation was not associated with response in any subset (**Figure S13E-F**). When combining the Lund and SWOG S1314 cohorts, proliferation was associated with better response only within the Uro subtype, with a stronger effect in the MVAC treated Uro tumors (p=0.0016) than in the GC treated Uro tumors (p=0.12) (**Figure S13G**). Notably, in non-Uro subtypes, proliferation showed no association with response in either regimen (**Figure S13H**). These data support a link between proliferation and treatment response in the Uro subtype that is more pronounced among patients that received MVAC compared to GC.

Next, resampling-based tests were undertaken to determine to what degree the observed meta rank-score distributions differed from what would be expected under the null hypothesis, i.e. if the bulk TUR-BT transcriptome carried no predictive information for NAC response. To this end, 100 permutations of the meta rank-score were calculated from the same three datasets, but with response labels randomly assigned. The extremes of these null distributions are plotted in **Figure 4C-F**. In the full cohort, the top 150 response-associated genes obtained more extreme meta rank-scores than 95% of the permuted distributions (**Figure 4C**). Subtype-specific analyses revealed that in the Uro subtype, more than 200 genes were more strongly associated with NAC response than expected by chance (**Figure 4D**). In contrast, the GU subtype showed no significant deviations from the null distribution, even among the genes with the lowest score (**Figure 4E**). Finally, in the Ba/Sq subtype, approximately 40 genes obtained more extreme scores than expected (**Figure 4F**). Notably, no significant differences were detected between the real and permuted meta rank-score distributions in the non-response direction in any cohort.

In summary, these results confirm that substantial predictive signals for NAC sensitivity are present in MIBC expression profiles, which can be attributed mainly to a few hundred genes in the Uro subtype and several dozens of genes in the Ba/Sq subtype. The lack of robust signals in the GU subtype, and in the direction of non-response, indicates that few, if any, genes are predictive of NAC response in these settings.

### Protein-level validation of candidate subtype-dependent biomarkers

Subsequently, we used the meta rank-scores to prioritize six candidate biomarkers for protein-level validation. In addition to an extreme meta rank-score in the NAC cohorts and not in the RC-only cohort, these biomarker candidates should have a commercially available antibody suitable for IHC and show a significant univariate association to cancer-specific survival in the Lund NAC cohort for which tumor tissue embedded in TMAs was available. Based on these criteria, we selected *SMCO4* and *CCNB1* (response in Uro), *CD55* (non-response in GU), *NKG7* (response in Ba/Sq), *ALDH1A1* (response in Uro and non-response in GU), and *CYP4Z1* (non-response in Uro) (**Table S1**). Adequate staining was achieved for four of the six candidate markers: CCNB1, CD55, NKG7, and ALDH1A1. IHC-scores correlated positively with corresponding gene expression values (Pearson *r* = 0.47-0.52) (**Figure S14**). IHC-scores stratified by response-status were then analyzed in the intended subsets. The density of CCNB1-positive cancer cells was associated with NAC response in the Uro subtype only (p≤0.0001) (**Figure 5B**). This association was not related to the chemotherapy regimen (**Figure S15**). CD55, which was expressed in both cancer and non-cancer cells, showed a non-significant trend toward non-response in the GU subtype, with no effect in the other subtypes (**Figure 5C**). No association with NAC response was observed for ALDH1A1 staining across molecular subtype or cellular compartment (cancer cells, non-cancer cells generally, or in stellate-shaped myeloid cells) (**Figure S16**). Finally, despite few responders in the Ba/Sq subtype, the density of NKG7-positive immune cells was significantly associated with NAC response in this subset (p=0.003) (**Figure 5D**). In the Lund RC-only cohort, no association with pCR was observed for CCNB1 or NKG7. For CD55, a moderate association was observed, but only in non-GU tumors and in the opposite direction of that seen in the NAC cohorts. A weak association with pCR status was also seen for ALDH1A1 in the Uro subtype (**Figure S17**).

**Figure 5.**
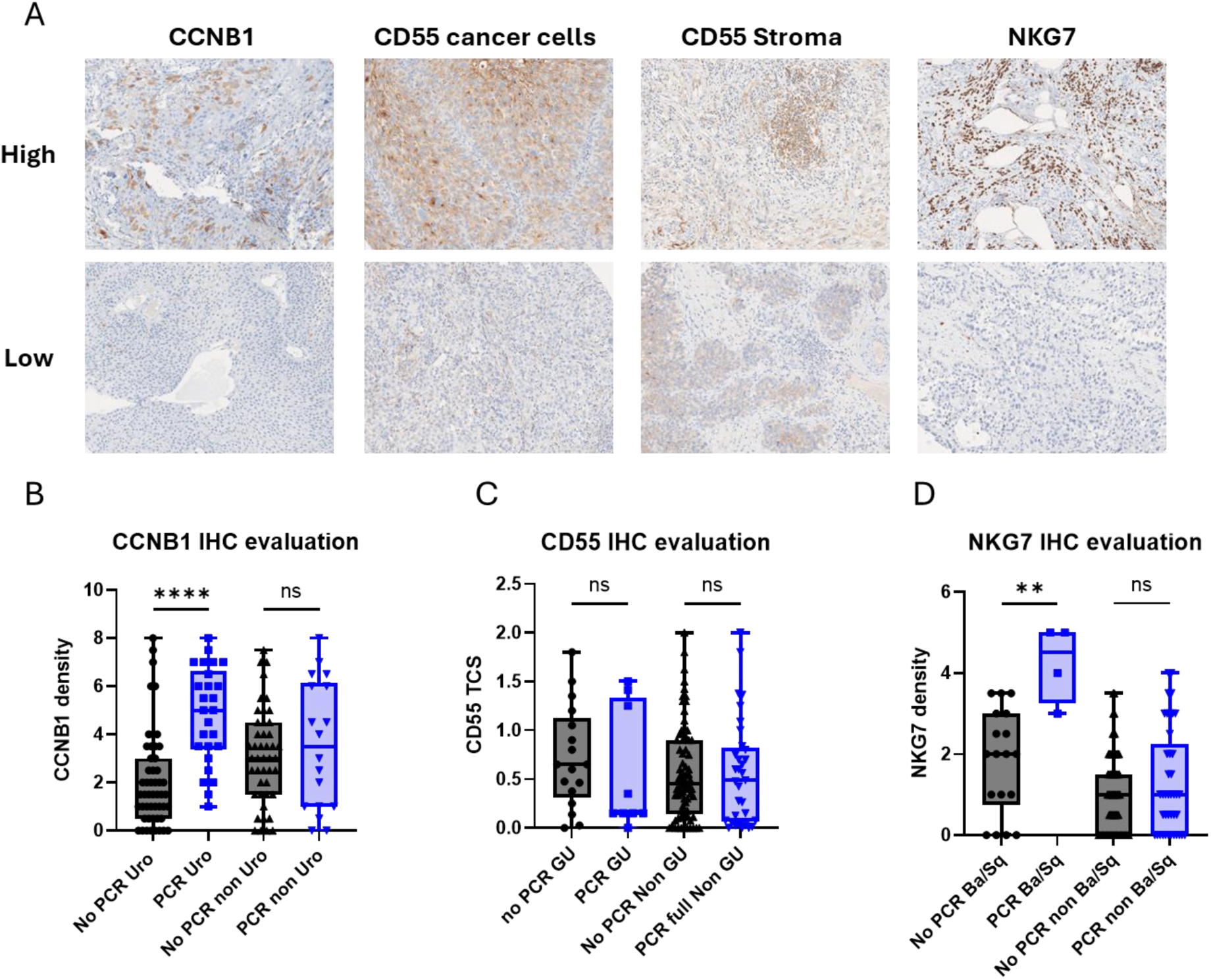
IHC evaluation of candidate biomarkers confirms their specific subtype effect on treatment sensitivity. A: The upper panels show strong protein staining in a TMA core from the Lund cohort, with high expression observed in tumor cells for all biomarkers and in the stroma for CD55. The lower panels show weak protein expression in the same TMA core. B-D: Protein expression was evaluated in cancer cells for CCNB1 and NKG7 and in both cancer cell and stroma compartments for CD55. Expression levels were then compared across tumor subtypes stratified by treatment response: Uro and non-Uro for CCNB1 (B), GU and non-GU for CD55 (C), and Ba/Sq and non-Ba/Sq for NKG7 (D).

In summary, IHC staining of CCNB1 and NKG7 protein expression captures the chemotherapy sensitivity associated with proliferation in the Uro subtype, and cytotoxic immune activity in the Ba/Sq subtypes, respectively.

## Discussion

The major finding of this work is that broad biological programs relating to proliferation and to cytotoxic T- and NK-cells are strongly associated with pathologic response, but in a subtype-dependent manner. Proliferation genes were the strongest predictors of response in the Urothelial-like (Uro) subtype, whereas cytotoxic immune signatures were predictive specifically in the Basal/Squamous (Ba/Sq) subtype. Notably, previous studies have reported positive associations between neoadjuvant therapy response and various measures of proliferation [16, 30, 31] as well as T-cell activity [9, 13, 14, 32, 33], but these findings were not previously linked to a specific molecular subtype. Galsky and colleagues showed in the IMvigor130 trial that pre-existing adaptive immunity, including T- and NK-effector signatures, was required for the improved effect of cisplatin over carboplatin [33]. Although the clinical setting with palliative treatment for locally advanced and metastatic disease is different from the current neoadjuvant context, this work highlights the role of cisplatin in enhancing tumor immunogenicity and promoting adaptive anti-tumor immunity. Similarly, Groeneveld *et al*, using data from the VESPER-trial, reported that tumors with mixed molecular subtype and low proliferation had poor pCR rate [11]. It remains to be seen if this is in line with our result, since tumors with a mixed subtype often involved the Stroma-rich MIBC-consensus subtype, which has a lower proliferation signal due to higher stromal content in bulk tissue samples. The LundTax classification avoids conflation of tissue sample purity with molecular subtype, and it would be interesting to apply proliferation and T- and NK-cell signatures in the light of LundTax classification to the IMvigor130 and VESPER datasets.

A major limitation in the search for predictive biomarkers of NAC response in MIBC is that no transcriptomic studies have randomized patients to NAC versus RC-only. Therefore, current evidence is limited to single-arm clinical trials and retrospective cohort studies, all hampered by difficulties to separate predictive effect from general prognostic associations. In this context, pCR is an important surrogate endpoint because survival after RC depends on several factors other than treatment response e.g. clinical T- and N-stage, and presence of lymphovascular invasion or a high-risk variant histology [23, 34]. However, the probability of obtaining a pathologic response in the bladder (pT0) does not only depend on the tumor’s intrinsic treatment sensitivity, but also on the initial disease burden and TUR-BT completeness. The contribution of TUR-BT to pCR rates in cT2 disease has been estimated at 38%, with pCR observed in 28% of patients receiving NAC compared to 9% without [35]. In non-organ confined disease, pCR rates are lower but NAC continues to confer a survival benefit [36]. Furthermore, differences in treatment regimens (GC or MVAC); inclusion of clinically node-positive patients despite being a separate induction indication for preoperative chemotherapy compared to NAC; the number of NAC cycles; response definitions (pT0N0 versus downstaging to pTis/Ta/T1); population-based versus selected cohorts; and diverse patient populations are examples of confounders that likely affect studies without a control arm not receiving NAC. Such variability may explain the low reproducibility of biomarkers reported in individual studies. In this work, we hypothesize that genes robustly associated with pathologic response should demonstrate consistent signals across multiple studies, even if they are not the top differentially expressed genes in each individual study. Furthermore, since MIBC molecular subtypes represent biologically distinct entities [25, 37], we propose that different subtypes may respond or fail to respond to NAC through distinct biological mechanisms and that subtype-specific predictive genes may be obscured in pan-MIBC investigations.

To mitigate these limitations, we integrated all available NAC-treated cohorts with transcriptomic data and sufficient sample size. By relying on gene ranking rather than absolute expression values, we minimize cohort-specific biases. We also applied uniform response definitions and inclusion criteria regarding number of received NAC cycles. Furthermore, by stratifying the analysis according to the cancer cells’ intrinsic phenotypic state with the LundTax classifier [24], we accounted for the biological heterogeneity of MIBC and could dissect NAC response biology in a subtype-informed manner. Thus, small scale studies, although underpowered for robust conclusions, can still be contextualized in light of our meta rank-scores. As an example, Murphy *et al*. reported DEGs in a single-center MIBC cohort (n=18), highlighting expression of *CRKL* in responders and *SPRY1* in non-responders [38]. Of these, only *SPRY1* showed a consistent non-response association in the Uro (rank 178) and Ba/Sq subsets, but, surprisingly the opposite association in GU (rank 205 response-associated). Given that only two clear GU/LumU tumors were included in their discovery cohort, subtype imbalance is a possible explanation for this discrepancy [38]. Thus, our result nuanced the interpretation to suggest that *SPRY1* could be a candidate subtype-dependent NAC biomarker warranting further investigation.

We selected six candidate markers to validate by IHC, of which two did not yield satisfactory staining (SMCO4 and CYP4Z1), one failed to reproduce the expected association (ALDH1A1), and one showed a non-significant trend consistent with the mRNA data (CD55). Statistically significant associations were confirmed for proliferation (CCNB1, Uro) and cytotoxic immunity (NKG7, Ba/Sq), both with effect sizes indicating potential clinical utility in the Lund NAC cohort. Although not an independent cohort validation, these findings show that our RNA-level observations reflect true biological processes, namely the density of proliferating cancer cells and intra-tumoral cytotoxic lymphocytes, rather than tissue sample purity effects or technical artifacts.

The main limitation of our work is its restriction to expressed biomarkers at the bulk transcriptomic level. Some determinants of NAC response may only become evident through temporally or spatially resolved data, by accounting for non-linear relationship between genes, or through integration with proteomic and genomic information. In this sense, our work captures only part of the context dependency of NAC response prediction. Nevertheless, central aspects of MIBC biology including molecular subtype, immune-infiltration and proliferation are well captured in the bulk transcriptome, and our results suggest that their predictive potential is now beginning to emerge. Another limitation is that our analysis was based on microarray data, whereas RNA-sequencing, the current technical standard, was not used for the suitable NAC cohorts. Still, by relying on internal gene-ranks within each cohort, our approach is inherently robust across platforms, meaning that as additional RNA-seq cohorts become available, they can be added to refine and expand the subtype-specific meta rank-scores.

In summary, we present the first subtype-informed, integrated analysis of NAC-response biomarkers across the largest publicly available transcriptomic cohorts. Our findings show that proliferation and cytotoxic T- and NK-cell signatures are predictive of NAC-response specifically in the Uro and Ba/Sq subtypes, respectively. The resulting meta rank-score tables (**Table S1**) provide a resource for biomarker discovery, which can be further strengthened as new transcriptomic data becomes available.

## Data availability

The data generated in this study are available within the article and its supplementary data files. Expression profile data from Lund cohort and Seiler2017 cohort analyzed in this study were obtained from Gene Expression Omnibus (GEO) at GSE169455, GSE83586, and GSE87304. The SWOG S1314 cohort data analysed in this study were provided by the Southwest Oncology Group. Restrictions apply to the availability of these data, which were used under license for this study.

## Author contributions

AZ and GS designed the project. PE, TWF, RS, CMT, PCB, SPL, and DMC provided molecular and patient data. AZ, GS, PE and CAM performed the cohort analyses. AZ and GS conducted data integration and generated the meta-rank score tables. AZ performed the signature analysis. AZ and PE carried out the resampling-based analysis. LT optimized and performed IHC staining. AZ, CB, and GS performed IHC evaluations. AZ and GS interpreted the results and wrote the manuscript. All authors reviewed and edited the manuscript.

## References

1. Advanced Bladder Cancer (ABC) Meta-analysis Collaboration. Neoadjuvant chemotherapy in invasive bladder cancer: update of a systematic review and meta-analysis of individual patient data advanced bladder cancer (ABC) meta-analysis collaboration. Eur Urol. 2005. Aug;48(2):202-5

2. Witjes JA, Bruins HM, Carrión A, Cathomas R, Compérat E, Efstathiou JA, et al. European Association of Urology Guidelines on Muscle-invasive and Metastatic Bladder Cancer: Summary of the 2023 Guidelines. Eur Urol. 2024 Jan;85(1):17–31.

3. Powles T, Catto JWF, Galsky MD, Al-Ahmadie H, Meeks JJ, Nishiyama H, et al. Perioperative Durvalumab with Neoadjuvant Chemotherapy in Operable Bladder Cancer. N Engl J Med. 2024 Nov 14;391(19):1773–1786.

4. Rosenblatt R, Sherif A, Rintala E, Wahlqvist R, Ullén A, Nilsson S, et al. Pathologic downstaging is a surrogate marker for efficacy and increased survival following neoadjuvant chemotherapy and radical cystectomy for muscle-invasive urothelial bladder cancer. Eur Urol. 2012 Jun;61(6):1229–38.

5. Lavery HJ, Stensland KD, Niegisch G, Albers P, Droller MJ. Pathological T0 following radical cystectomy with or without neoadjuvant chemotherapy: a useful surrogate. J Urol. 2014 Apr;191(4):898–906.

6. Petrelli F, Coinu A, Cabiddu M, Ghilardi M, Vavassori I, Barni S. Correlation of pathologic complete response with survival after neoadjuvant chemotherapy in bladder cancer treated with cystectomy: a meta-analysis. Eur Urol. 2014 Feb;65(2):350–7.

7. Bhindi B, Frank I, Mason RJ, Tarrell RF, Thapa P, Cheville JC, et al. Oncologic Outcomes for Patients with Residual Cancer at Cystectomy Following Neoadjuvant Chemotherapy: A Pathologic Stage-matched Analysis. Eur Urol. 2017 Nov;72(5):660–664.

8. Reesink DJ, Voskuilen CS, van de Garde EMW, Hendricksen K, Horenblas S, van Melick HHE, et al. Survival in Responders and Nonresponders of Neoadjuvant and Induction Chemotherapy in Invasive Urothelial Carcinoma of the Urinary Bladder: A Clinical and Pathological Stage-Matched Analysis. Clin Genitourin Cancer. 2025 Feb 21:102319.

9. Taber A, Christensen E, Lamy P, Nordentoft I, Prip F, Lindskrog SV, et al. Molecular correlates of cisplatin-based chemotherapy response in muscle invasive bladder cancer by integrated multi-omics analysis. Nat Commun. 2020 Sep 25;11(1):4858.

10. Sjödahl G, Abrahamsson J, Holmsten K, Bernardo C, Chebil G, Eriksson P, et al. Different Responses to Neoadjuvant Chemotherapy in Urothelial Carcinoma Molecular Subtypes. Eur Urol. 2022 May;81(5):523–532.

11. Groeneveld CS, Pfister C, Culine S, Harter V, Krucker C, Fontugne J, et al. Basal/squamous and mixed subtype bladder cancers present poor outcomes after neoadjuvant chemotherapy in the VESPER trial. Ann Oncol. 2025 Jan;36(1):89–98.

12. Börcsök J, Gopaul D, Devesa-Serrano D, Mooser C, Jonsson N, Cagiada M, et al. Quantitative functional profiling of ERCC2 mutations deciphers cisplatin sensitivity in bladder cancer.J Clin Invest. 2025 Aug 15;135(16):e186688.

13. Vollmer T, Schlickeiser S, Amini L, Schulenberg S, Wendering DJ, Banday V, et al. The intratumoral CXCR3 chemokine system is predictive of chemotherapy response in human bladder cancer. Sci Transl Med. 2021 Jan 13;13(576):eabb3735.

14. Nassif EF, Mlecnik B, Thibault C, Auvray M, Bruni D, Colau A, et al. The Immunoscore in Localized Urothelial Carcinoma Treated with Neoadjuvant Chemotherapy: Clinical Significance for Pathologic Responses and Overall Survival. Cancers (Basel). 2021 Jan 28;13(3):494.

15. Choi W, Porten S, Kim S, Willis D, Plimack ER, Hoffman-Censits J, et al. Identification of distinct basal and luminal subtypes of muscle-invasive bladder cancer with different sensitivities to frontline chemotherapy. Cancer Cell. 2014 Feb 10;25(2):152–65.

16. Mi H, Bivalacqua TJ, Kates M, Seiler R, Black PC, Popel AS, et al. Predictive models of response to neoadjuvant chemotherapy in muscle-invasive bladder cancer using nuclear morphology and tissue architecture. Cell Rep Med. 2021 Aug 27;2(9):100382.

17. Liu D, Plimack ER, Hoffman-Censits J, Garraway LA, Bellmunt J, Van Allen E, et al. Clinical Validation of Chemotherapy Response Biomarker ERCC2 in Muscle-Invasive Urothelial Bladder Carcinoma. JAMA Oncol. 2016 Aug 1;2(8):1094–6.

18. Gil-Jimenez A, van Dorp J, Contreras-Sanz A, van der Vos K, Vis DJ, Braaf L, et al. Assessment of Predictive Genomic Biomarkers for Response to Cisplatin-based Neoadjuvant Chemotherapy in Bladder Cancer. Eur Urol. 2023 Apr;83(4):313–317.

19. Flaig TW, Tangen CM, Daneshmand S, Alva A, Lerner SP, Lucia MS, et al. A Randomized Phase II Study of Coexpression Extrapolation (COXEN) with Neoadjuvant Chemotherapy for Bladder Cancer (SWOG S1314; NCT02177695). Clin Cancer Res. 2021 May 1;27(9):2435–2441.

20. Lerner SP, McConkey DJ, Tangen CM, Meeks JJ, Flaig TW, Hua X, et al. Association of Molecular Subtypes with Pathologic Response, PFS, and OS in a Phase II Study of COXEN with Neoadjuvant Chemotherapy for Muscle-invasive Bladder Cancer. Clin Cancer Res. 2024 Jan 17;30(2):444–449

21. Seiler R, Ashab HAD, Erho N, van Rhijn BWG, Winters B, Douglas J, et al. Impact of Molecular Subtypes in Muscle-invasive Bladder Cancer on Predicting Response and Survival after Neoadjuvant Chemotherapy. Eur Urol. 2017 Oct;72(4):544–554.

22. Sjödahl G, Eriksson P, Liedberg F, Höglund M. Molecular classification of urothelial carcinoma: global mRNA classification versus tumour-cell phenotype classification. J Pathol. 2017 May;242(1):113–125.

23. Kollberg P, Chebil G, Eriksson P, Sjödahl G, Liedberg F. Molecular subtypes applied to a population-based modern cystectomy series do not predict cancer-specific survival. Urol Oncol. 2019 Oct;37(10):791–799.

24. Cotillas EA, Bernardo C, Veerla S, Liedberg F, Sjödahl G, Eriksson P. A Versatile and Upgraded Version of the LundTax Classification Algorithm Applied to Independent Cohorts. J Mol Diagn. 2024 Dec;26(12):1081–1101.

25. Kamoun A, de Reyniès A, Allory Y, Sjödahl G, Robertson AG, Seiler R, et al. A Consensus Molecular Classification of Muscle-invasive Bladder Cancer. Eur Urol. 2020 Apr;77(4):420–433.

26. Ge SX, Jung D, Yao R. ShinyGO: a graphical gene-set enrichment tool for animals and plants. Bioinformatics. 2020 Apr 15;36(8):2628–2629.

27. Subramanian A, Tamayo P, Mootha VK, Mukherjee S, Ebert BL, Gillette MA, et al. Gene set enrichment analysis: a knowledge-based approach for interpreting genome-wide expression profiles. Proc Natl Acad Sci U S A. 2005 Oct 25;102(43):15545–50.

28. Doncheva NT, Morris JH, Gorodkin J, Jensen LJ. Cytoscape StringApp: Network Analysis and Visualization of Proteomics Data. J Proteome Res. 2019 Feb 1;18(2):623–632.

29. Khurana S, George SP. Regulation of cell structure and function by actin-binding proteins: villin’s perspective. FEBS Lett. 2008 Jun 18;582(14):2128–39.

30. Fleischmann A, Thalmann GN, Perren A, Seiler R. Tumor regression grade of urothelial bladder cancer after neoadjuvant chemotherapy: a novel and successful strategy to predict survival. Am J Surg Pathol. 2014 Mar;38(3):325–32.

31. Beckabir W, Wobker SE, Damrauer JS, Midkiff B, De la Cruz G, Makarov V, et al. Spatial Relationships in the Tumor Microenvironment Demonstrate Association with Pathologic Response to Neoadjuvant Chemoimmunotherapy in Muscle-invasive Bladder Cancer. Eur Urol. 2024 Mar;85(3):242–253.

32. Baras AS, Drake C, Liu JJ, Gandhi N, Kates M, Hoque MO, et al. The ratio of CD8 to Treg tumor-infiltrating lymphocytes is associated with response to cisplatin-based neoadjuvant chemotherapy in patients with muscle invasive urothelial carcinoma of the bladder. Oncoimmunology. 2016 Feb 18;5(5):e1134412.

33. Galsky MD, Guan X, Rishipathak D, Rapaport AS, Shehata HM, Banchereau R, et al. Immunomodulatory effects and improved outcomes with cisplatin- versus carboplatin-based chemotherapy plus atezolizumab in urothelial cancer. Cell Rep Med. 2024 Feb 20;5(2):101393.

34. Stein JP, Lieskovsky G, Cote R, Groshen S, Feng AC, Boyd S, et al. Radical Cystectomy in the Treatment of Invasive Bladder Cancer: Long-Term Results in 1,054 Patients. J Clin Oncol. 2023 Aug 1;41(22):3772–3781.

35. Brant A, Kates M, Chappidi MR, Patel HD, Sopko NA, Netto GJ, et al. Pathologic response in patients receiving neoadjuvant chemotherapy for muscle-invasive bladder cancer: Is therapeutic effect owing to chemotherapy or TURBT? Urol Oncol. 2017 Jan;35(1):34.e17-34.e25.

36. de Angelis M, Jannello LMI, Siech C, Baudo A, Di Bello F, Goyal JA, et al. Neoadjuvant chemotherapy before radical cystectomy in patients with organ-confined and non-organ-confined urothelial carcinoma. Urol Oncol. 2025 Jan;43(1):62.e1-62.e6.

37. Höglund M. What is a Bladder Cancer Molecular Subtype? Bladder Cancer. 2023 Dec 13;9(4):293–298.

38. Murphy N, Shih AJ, Shah P, Yaskiv O, Khalili H, Liew A, et al. Predictive molecular biomarkers for determining neoadjuvant chemosensitivity in muscle invasive bladder cancer. Oncotarget. 2022 Nov 2;13:1188–1200.

