## Supplementary material for "Proliferation- and cytotoxic immune signatures identify chemotherapy-responsive bladder tumors in a molecular subtype dependent manner": Figure S1-17 and Table S3

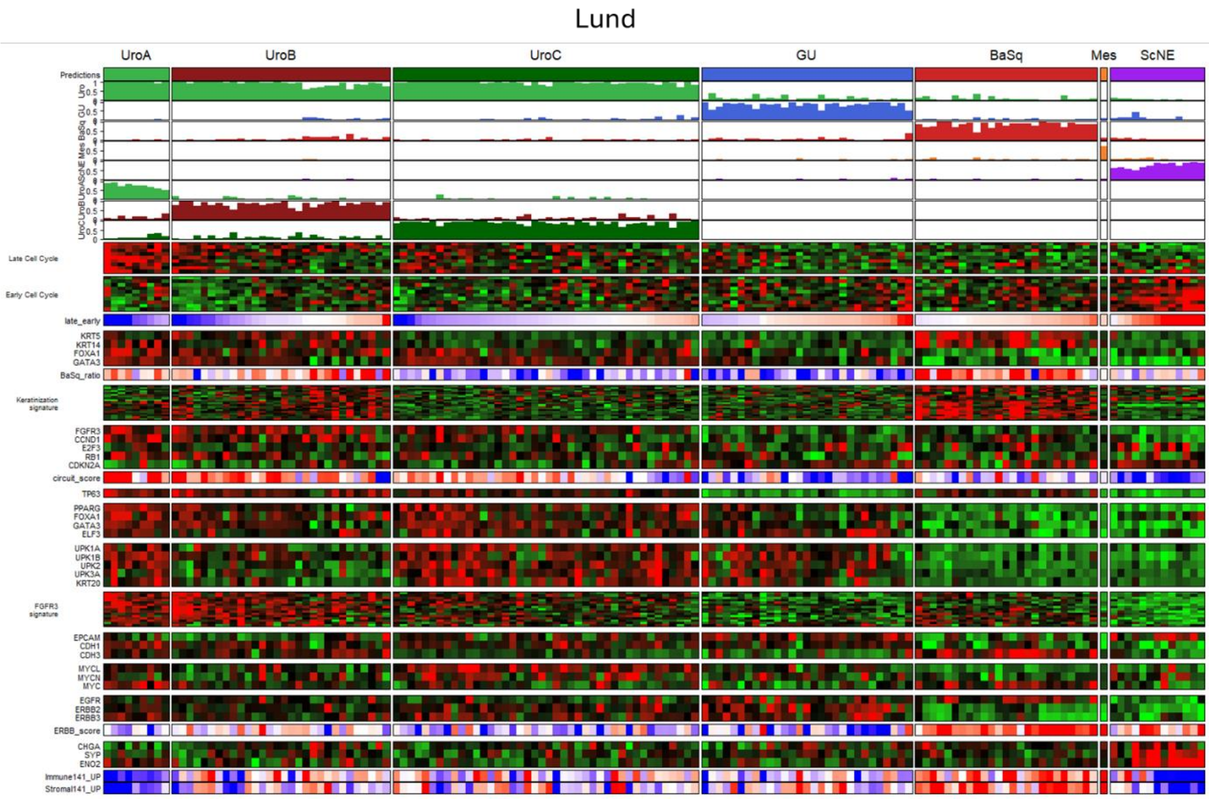


**Figure S1. Heatmap showing the subtype classification of the Lund NAC cohort**

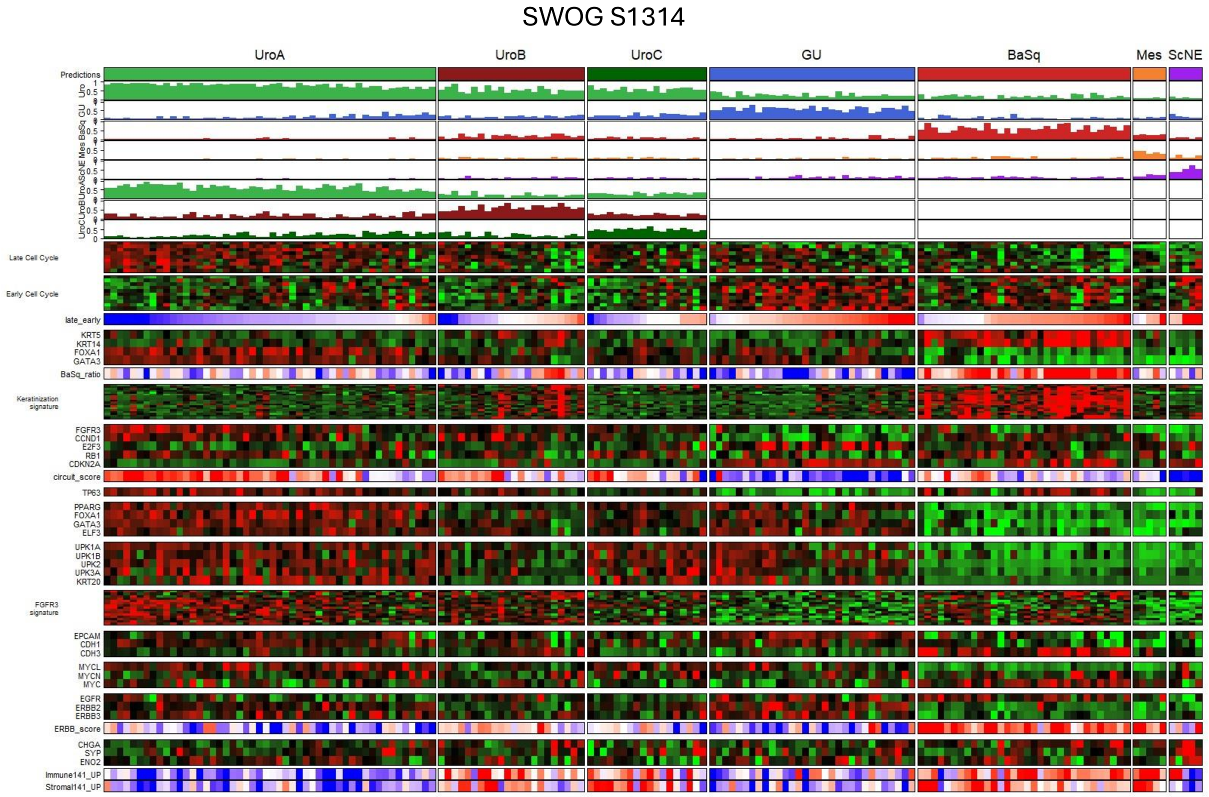


**Figure S2. Heatmap showing the subtype classification of the SWOG S1314 cohort**

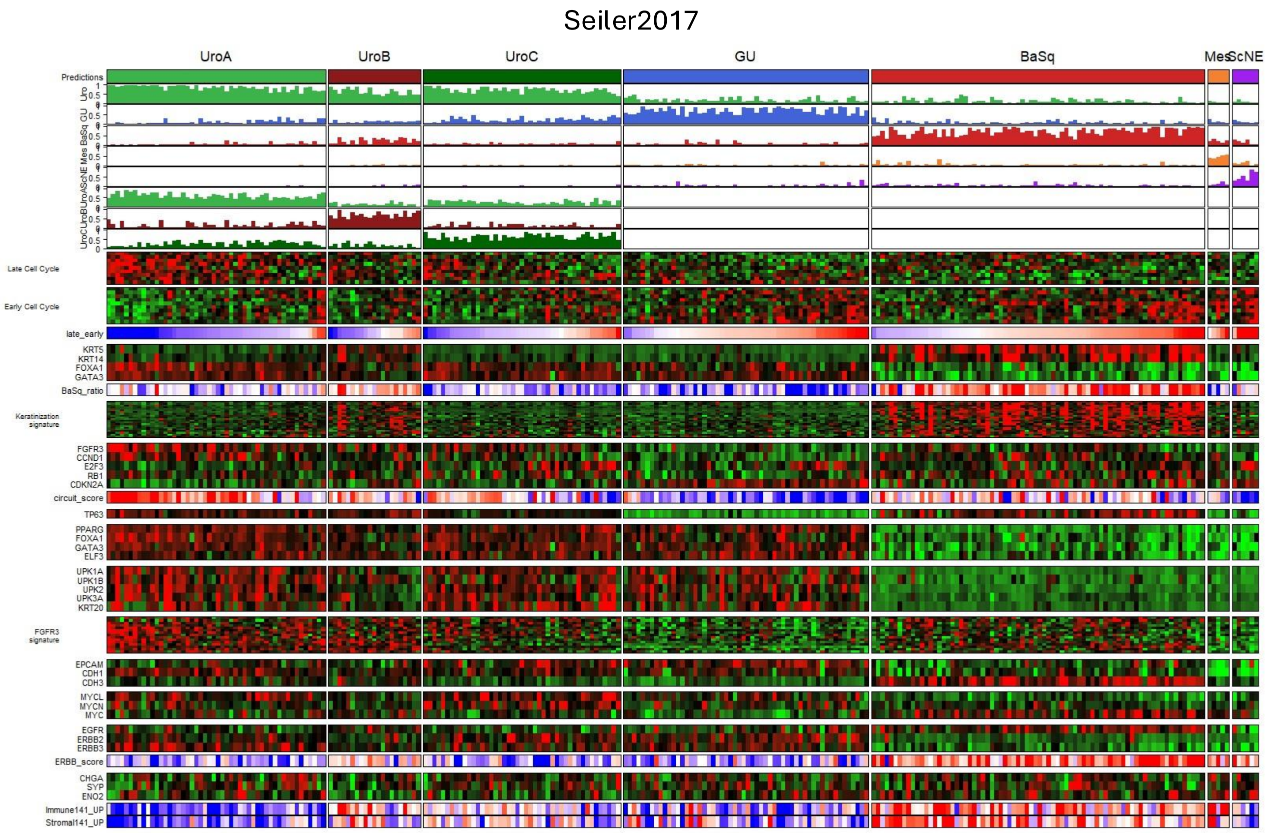


**Figure S3. Heatmap showing the subtype classification of the Seiler2017 cohort**

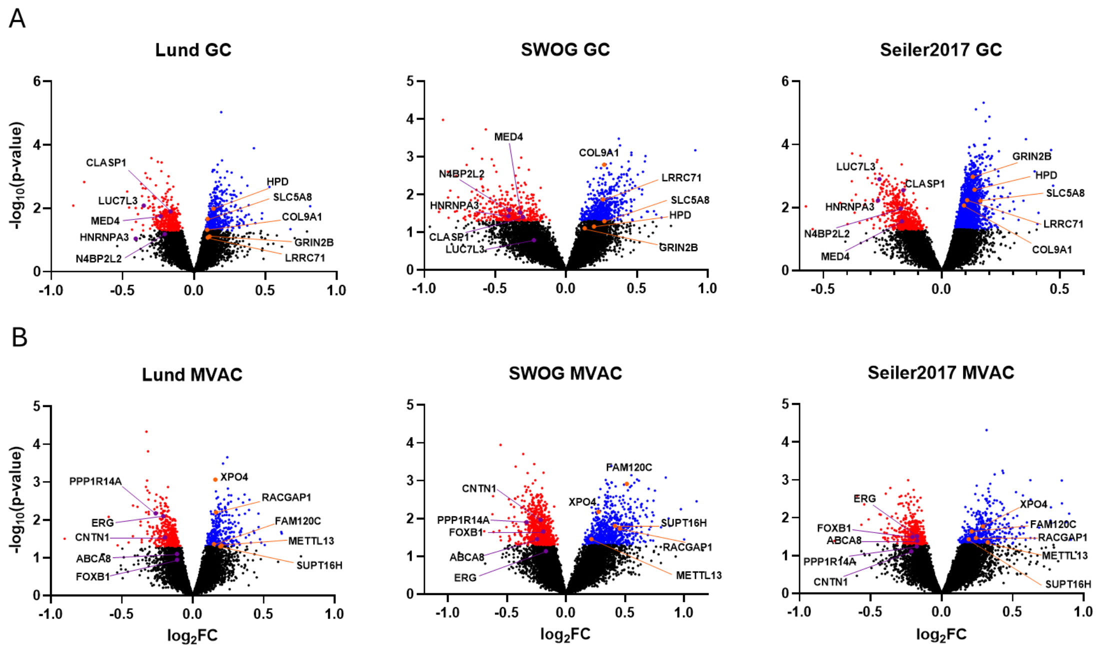


**Figure S4. Differential expression analysis by treatment response in each cohort stratified by treatment regimen.** *A-B. Volcano plots representing the differential expression analyses performed on the MVAC-treated subgroup (A) and GC-treated subgroup (B). Genes with statistical significance (p≤0.05) are marked in red for downregulation and blue for upregulation, and the 5 genes with the highest and lowest meta rank-score in each volcano plot are highlighted.*

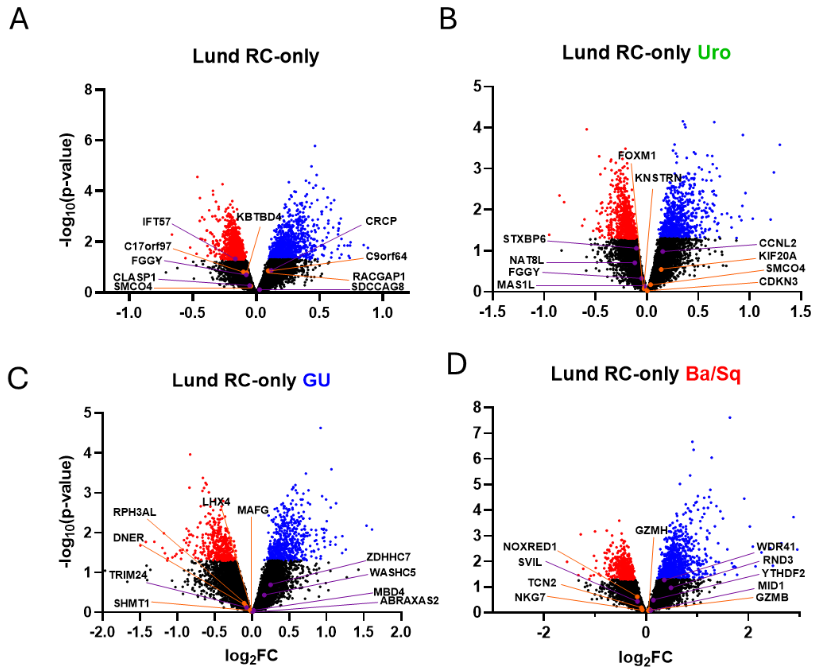


**Figure S5. Differential expression analysis by treatment response in the full Lund RC-only cohort and in three molecular subtypes.** *A-D. Volcano plots showing differentially expressed genes the full Lund RC-only cohort (A) and in the Uro (B), GU (C) and Ba/Sq (D) subsets. Statistically significant genes are marked in red for downregulation and blue for upregulation, and the 5 genes with the highest and lowest meta rank-score in each volcano plot are highlighted.*

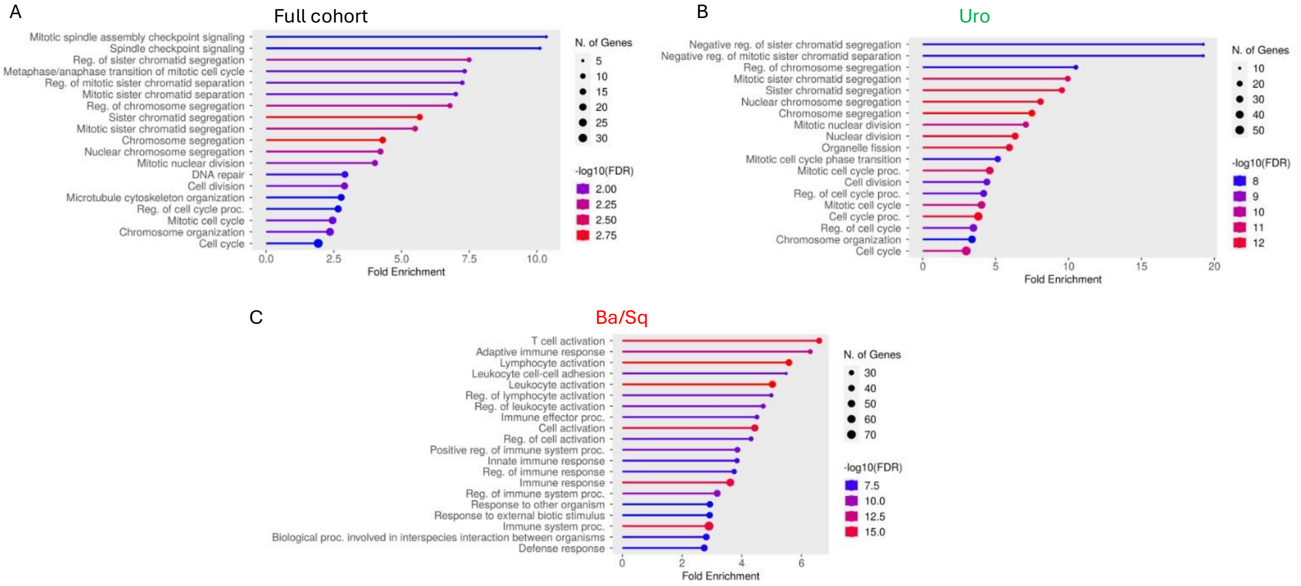


**Figure S6. Top 20 subtype-specific, response associated GO terms.** *GO term analysis using 'biological processes', for the 1% of genes with the lowest meta rank-score in the full cohort (A) and major molecular subtypes: Uro (B) and Ba/Sq (C). GU subtype, which had no significantly enriched processes in the top 1% of genes, is not shown.*

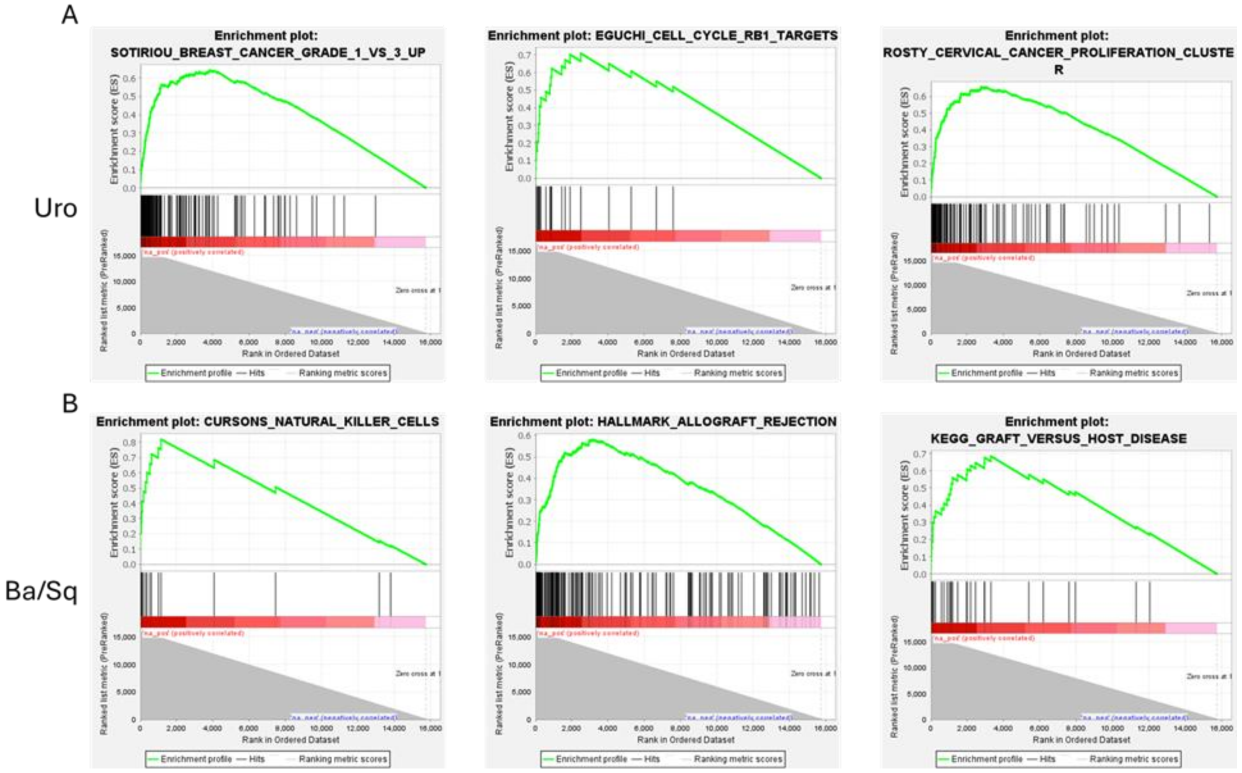


**Figure S7. The GSEA highlighted enrichments specific to molecular subtype.** *A and B: GSEA analysis confirmed robust enrichment of cell cycle-related signatures in the Uro subtype (A), and cytotoxic immune-related signatures in the Ba/Sq subtype (B), both positively associated with pathologic response.*

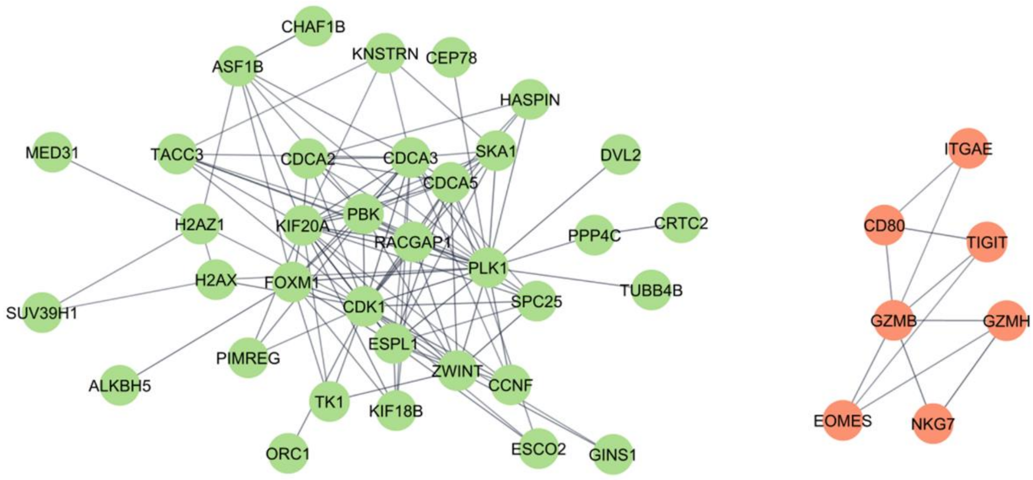


***Figure S8. The Uro subtype drives the protein interactome in the full cohort.*** *Major protein-protein interaction observed in the top 1% of genes with the lowest meta rank-score in the full cohort using STRING. The proliferation module enriched in the Uro subtype is identified, as well as a limited cytotoxic immune module enriched in Ba/Sq.*

A

**Lund cohort**

| Lund\consensus | LumP | LumNS | LumU | Ba/Sq | Stroma-rich | NE-like | Total |
|---|---|---|---|---|---|---|---|
| Uro | 39 | 18 | 7 | 7 | 10 | 0 | 81 |
| GU | 1 | 2 | 12 | 2 | 12 | 0 | 29 |
| Ba/Sq | 0 | 0 | 0 | 22 | 3 | 0 | 25 |
| ScNE | 0 | 0 | 0 | 0 | 4 | 9 | 13 |
| Mes | 0 | 0 | 0 | 0 | 1 | 0 | 1 |
| Total | 40 | 20 | 19 | 31 | 30 | 9 | 149 |

| Lund\consensus | LumP | LumNS | LumU | Ba/Sq | Stroma-rich | NE-like | Total |
|---|---|---|---|---|---|---|---|
| UroA | 9 | 0 | 0 | 0 | 0 | 0 | 9 |
| UroB | 18 | 1 | 0 | 7 | 4 | 0 | 30 |
| UroC | 12 | 17 | 7 | 0 | 6 | 0 | 42 |
| GU | 1 | 2 | 12 | 2 | 12 | 0 | 29 |
| Ba/Sq | 0 | 0 | 0 | 22 | 3 | 0 | 25 |
| ScNE | 0 | 0 | 0 | 0 | 4 | 9 | 13 |
| Mes | 0 | 0 | 0 | 0 | 1 | 0 | 1 |
| Total | 40 | 20 | 19 | 31 | 30 | 9 | 149 |

B

**Seiler2017**

| Lund\consensus | LumP | LumNS | LumU | Ba/Sq | Stroma-rich | NE-like | Total |
|---|---|---|---|---|---|---|---|
| Uro | 44 | 34 | 17 | 9 | 11 | 0 | 115 |
| GU | 3 | 6 | 32 | 1 | 15 | 0 | 57 |
| Ba/Sq | 0 | 0 | 0 | 67 | 9 | 0 | 76 |
| ScNE | 0 | 0 | 0 | 3 | 0 | 3 | 6 |
| Mes | 0 | 0 | 0 | 4 | 1 | 0 | 5 |
| Total | 47 | 40 | 49 | 84 | 36 | 3 | 259 |

| Lund\consensus | LumP | LumNS | LumU | Ba/Sq | Stroma-rich | NE-like | Total |
|---|---|---|---|---|---|---|---|
| UroA | 32 | 14 | 2 | 0 | 2 | 0 | 50 |
| UroB | 8 | 1 | 0 | 9 | 3 | 0 | 21 |
| UroC | 4 | 19 | 15 | 0 | 6 | 0 | 44 |
| GU | 3 | 6 | 32 | 1 | 15 | 0 | 57 |
| Ba/Sq | 0 | 0 | 0 | 67 | 9 | 0 | 76 |
| ScNE | 0 | 0 | 0 | 3 | 0 | 3 | 6 |
| Mes | 0 | 0 | 0 | 4 | 1 | 0 | 5 |
| Total | 47 | 40 | 49 | 84 | 36 | 3 | 259 |

C

**SWOG S1314**

| Lund\Consensus | LumP | LumNS | LumU | Ba/Sq | Stroma-rich | NE-like | Total |
|---|---|---|---|---|---|---|---|
| Uro | 45 | 21 | 5 | 9 | 10 | 0 | 90 |
| GU | 3 | 2 | 17 | 4 | 5 | 0 | 31 |
| Ba/Sq | 0 | 0 | 0 | 32 | 0 | 0 | 32 |
| ScNE | 0 | 0 | 0 | 3 | 0 | 2 | 5 |
| Mes | 0 | 0 | 0 | 4 | 1 | 0 | 5 |
| Total | 48 | 23 | 22 | 52 | 16 | 2 | 163 |

| Lund\consensus | LumP | LumNS | LumU | Ba/Sq | Stroma-rich | NE-like | Total |
|---|---|---|---|---|---|---|---|
| UroA | 39 | 9 | 2 | 0 | 0 | 0 | 50 |
| UroB | 4 | 4 | 0 | 9 | 5 | 0 | 22 |
| UroC | 2 | 8 | 3 | 0 | 5 | 0 | 18 |
| GU | 3 | 2 | 17 | 4 | 5 | 0 | 31 |
| Ba/Sq | 0 | 0 | 0 | 32 | 0 | 0 | 32 |
| ScNE | 0 | 0 | 0 | 3 | 0 | 2 | 5 |
| Mes | 0 | 0 | 0 | 4 | 1 | 0 | 5 |
| Total | 48 | 23 | 22 | 52 | 16 | 2 | 163 |

***Figure S9. Concordance between the LundTax and the consensus classifier.*** *The closest subtypes between the LundTax and consensus classifications are highlighted in green.*

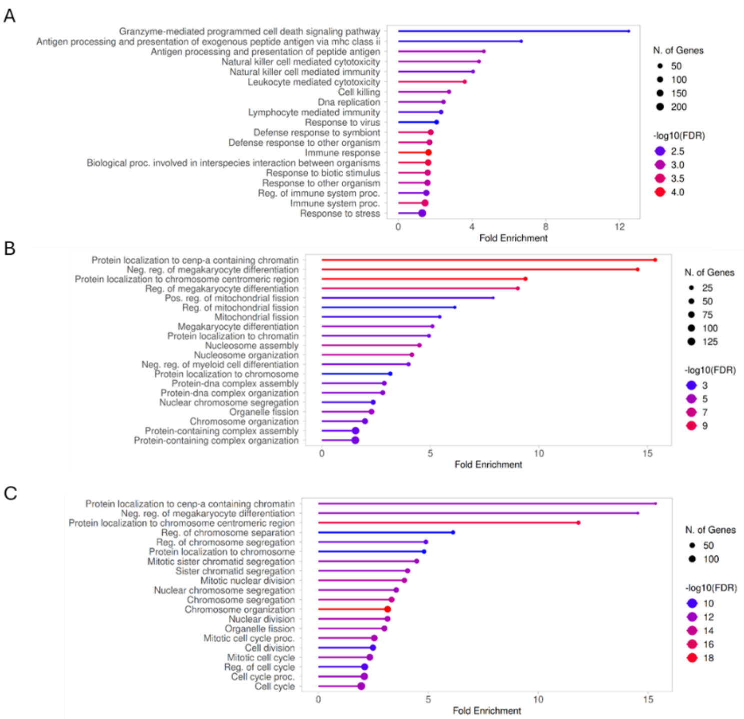


**Figure S10. Top 20 consensus subtype-specific, response associated GO terms.** *GO term analysis using 'biological processes', for the 5% of genes with the lowest meta rank-score in the consensus subtypes: Ba/Sq (A), LumNS (B) and the combination of LumNS and LumP (C). Stroma rich and LumU showed no enriched processes in the top 1% of genes.*

**Table S3. Pairwise overlap of top differentially expressed genes in the three NAC cohorts.**

| Subset | Top100 Responsive | Top100 Resistant | Top500 Responsive | Top500 Resistant |
|---|---|---|---|---|
| **All subtypes** | | | | |
| Lund : Seiler2017 | 4 | 0 | 27 | 12 |
| Lund : SWOG S1314 | 2 | 1 | 28 | 31 |
| Seiler2017 : SWOG S1314 | 0 | 2 | 24 | 16 |
| **Uro** | | | | |
| Lund : Seiler2017 | 1 | 1 | 23 (8 proliferation related) | 24 |
| Lund : SWOG S1314 | 13 | 2 | 79 (40 proliferation related) | 24 |
| Seiler2017 : SWOG S1314 | 1 | 1 | 28 (6 proliferation related) | 14 |
| **GU** | | | | |
| Lund : Seiler2017 | 1 | 4 | 27 | 29 |
| Lund : SWOG S1314 | 1 | 2 | 17 | 16 |
| Seiler2017 : SWOG S1314 | 0 | 0 | 16 | 13 |
| **Ba/Sq** | | | | |
| Lund : Seiler2017 | 2 | 2 | 36 (19 immune related) | 21 |
| Lund : SWOG S1314 | 1 | 1 | 17 (5 immune related) | 13 |
| Seiler2017 : SWOG S1314 | 2 | 1 | 16 (2 immune related) | 19 |

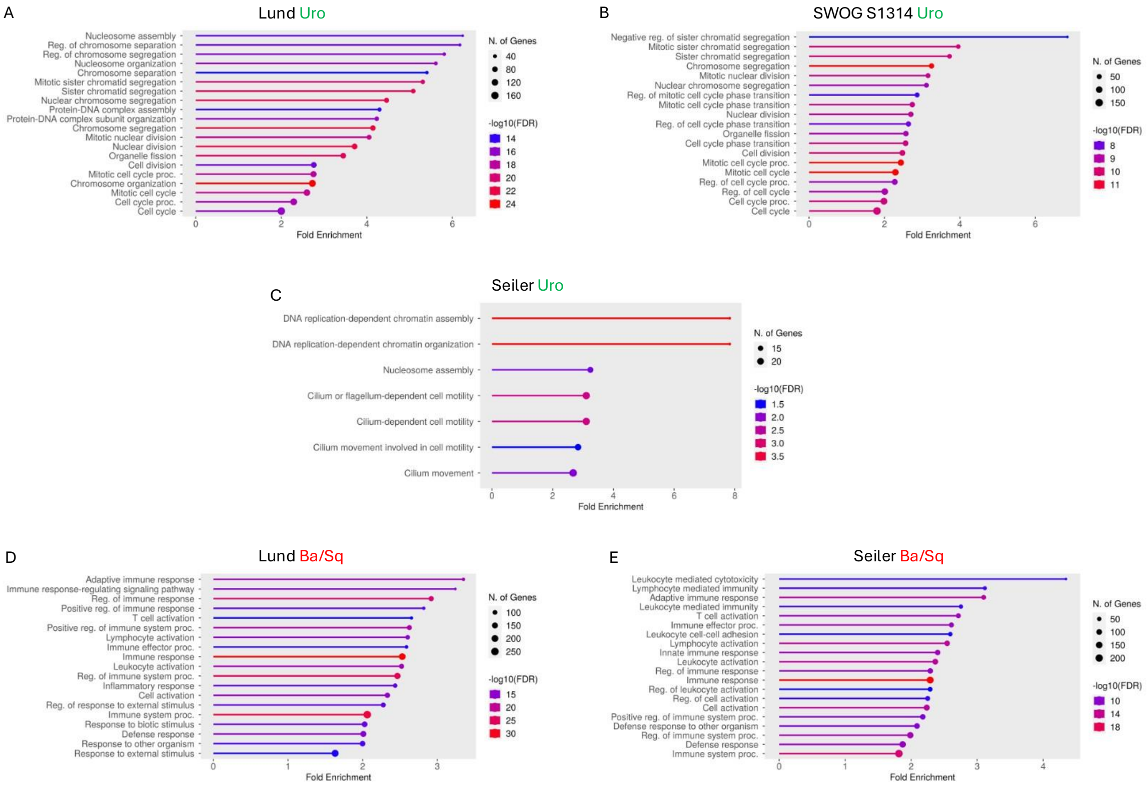


**Figure S11. Enrichment analyses are improved by combining the three cohorts.** *GO term analysis using 'biological processes', for the 5% most response-associated genes in the Uro and Ba/Sq subsets in each cohort. The Ba/Sq subtype in the SWOG S1314 cohort showed no significantly enriched processes.*

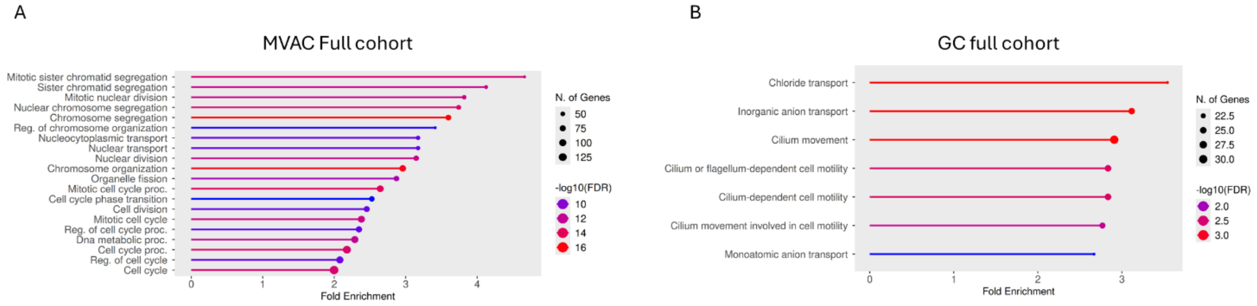


**Figure S12. Proliferation-related pathway enrichment is predominantly observed in patients treated with MVAC.** *GO term analysis using 'biological processes', for the 5% genes with the lowest meta rank-score. Analyses were conducted in the full cohort stratified by treatment regimen: MVAC (A) vs GC (B).*

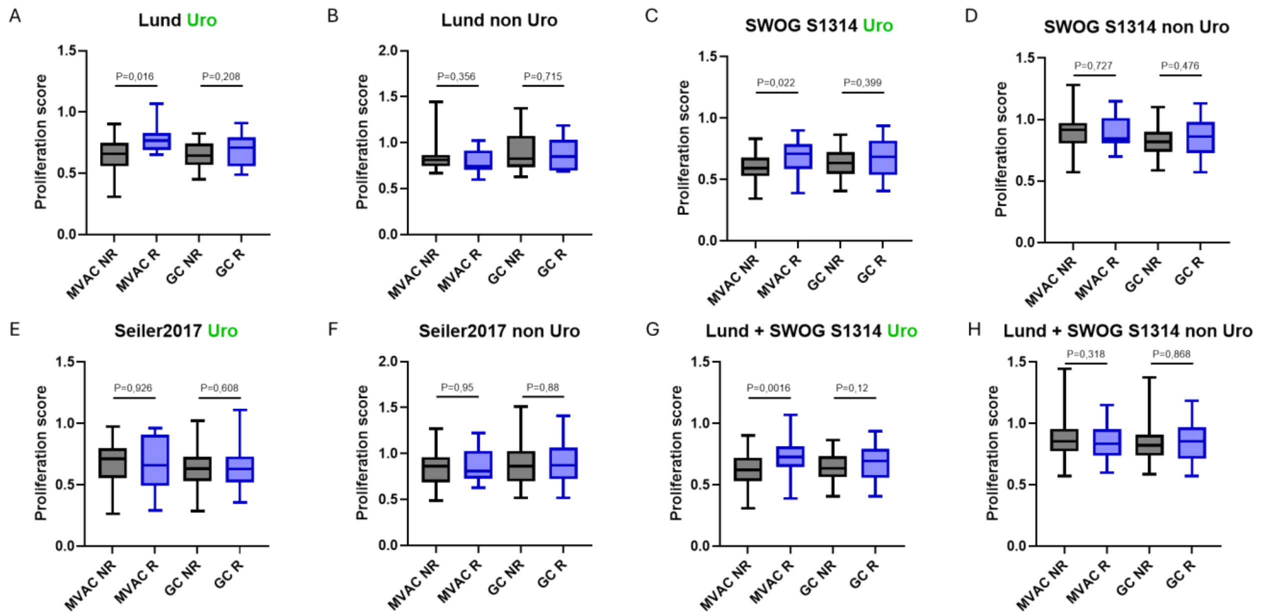


**Figure S13. Proliferation score is associated with response in the Uro subtype.** *A-H: Proliferation scores were computed for each patient and compared based on treatment response and treatment regimen within the Uro and non-Uro subtypes for the Lund cohort (A and B), SWOG (C and D), Seiler 2017 (E and F) and the combination of Lund and SWOG 2017 (G and H).*

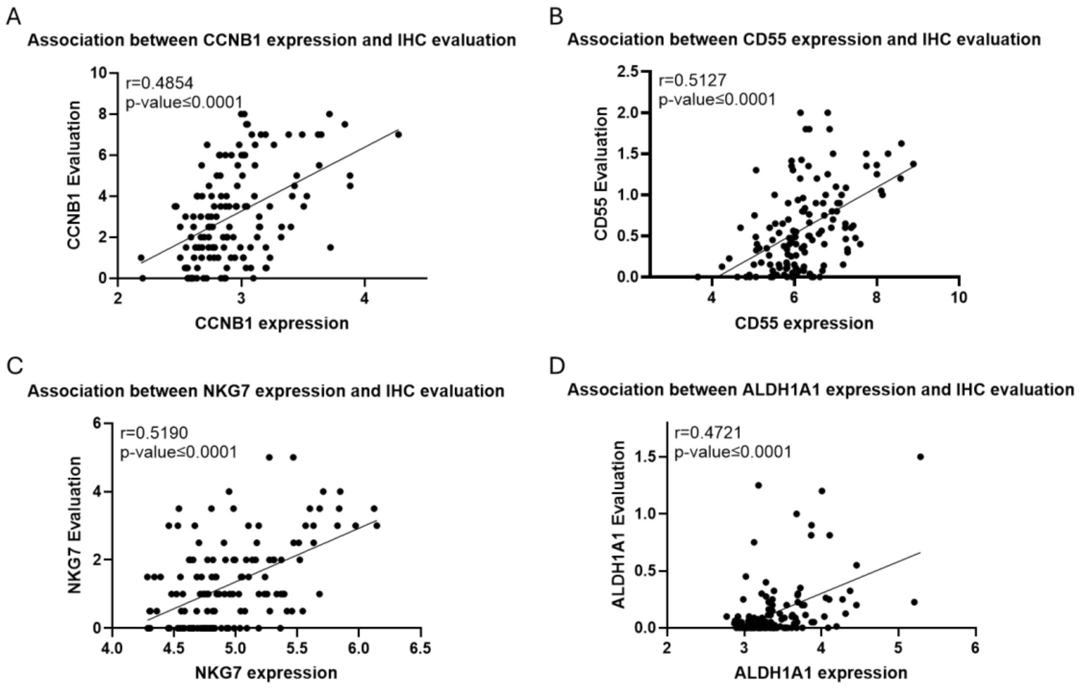


**Figure S14. The IHC evaluation of different biomarkers correlates with their gene expression.** *Scatter plots showing a significant positive correlation ($p \leq 0.0001$) between IHC evaluations from TMAs and matched mRNA expression for CCNB1 (A), CD55 (B), NKG7 (C), and ALDH1A1 (D) in the Lund NAC cohort.*

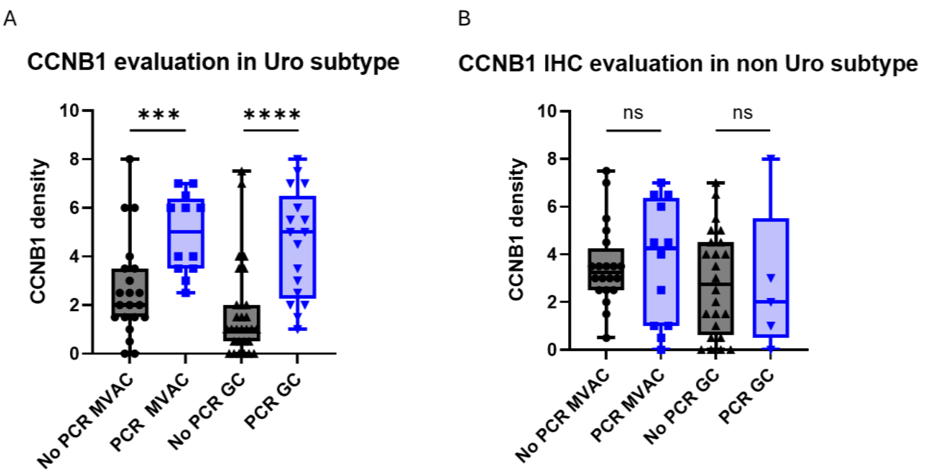


**Figure S15. The association between IHC evaluation of CCNB1 and response in Uro subtype is independent of the treatment regimen.** *A-B: CCNB1 expression was evaluated in cancer cells. Expression levels were then compared across tumor subtypes stratified by both treatment response and treatment regimen: Uro tumors treated with MVAC or GC (A) and non Uro tumors treated with MVAC or GC (B).*

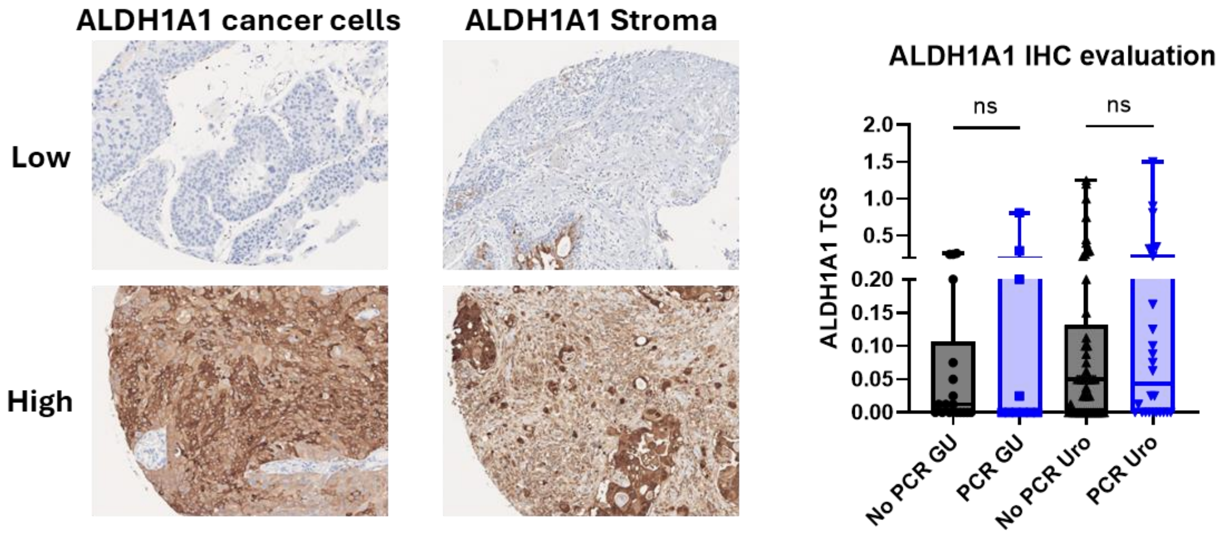


**Figure S16. IHC evaluation of ALDH1A1 does not confirm its potential role as a biomarker.** *The upper panels show examples of absent/weak ALDH1A1 staining in the cancer cells and in the stroma. The lower panels show examples with the strongest staining in the cancer cells and in the stroma. Boxplots show combined staining scores in the Uro and GU subtypes stratified by pathologic response.*

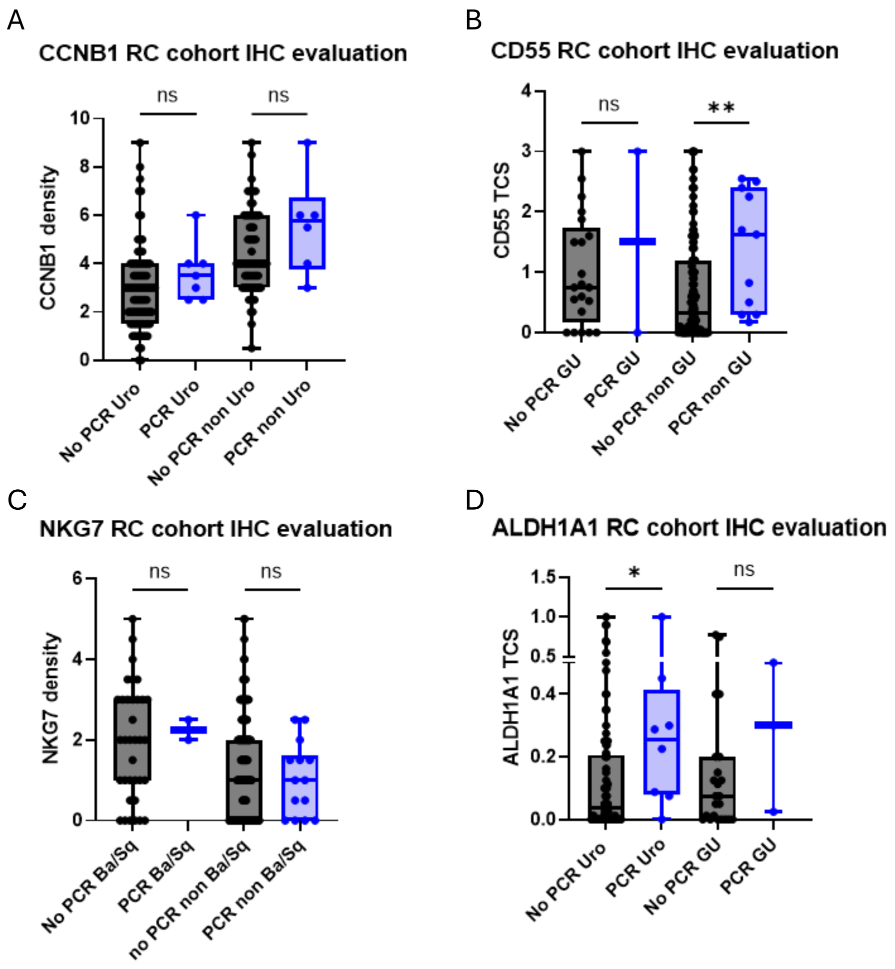


**Figure S17. NKG7 and CCNB1 serve as biomarkers of therapeutic response, but not as prognostic markers.** Density of *CCNB1 positive cancer cells, density of NKG7 positive immune cells, and quantitative expression in both cancer cell and stroma compartments for CD55 and ALH1A1 is shown in the LUND RC-only cohort. Analyses are stratified by each marker's subtype-specific association patterns and by pathologic response: Uro versus non-Uro for CCNB1 (A), GU versus non-GU for CD55 (B), Ba/Sq versus non-Ba/Sq for NKG7 (C) and Uro versus GU for ALDH1A1 (D).*
